# Systematic investigation of allelic regulatory activity of schizophrenia-associated common variants

**DOI:** 10.1101/2022.09.15.22279954

**Authors:** Jessica C. McAfee, Sool Lee, Jiseok Lee, Jessica L. Bell, Oleh Krupa, Jessica Davis, Kimberly Insigne, Marielle L. Bond, Douglas H. Phanstiel, Michael I. Love, Jason L. Stein, Sriram Kosuri, Hyejung Won

**Affiliations:** Department of Genetics, University of North Carolina at Chapel Hill, Chapel Hill, NC, 27599, USA; Neuroscience Center, University of North Carolina at Chapel Hill, Chapel Hill, NC, 27599, USA; Curriculum in Genetics and Molecular Biology, University of North Carolina, Chapel Hill, NC 27599, USA; Curriculum in Bioinformatics and Computational Biology, University of North Carolina at Chapel Hill, NC, 27599, USA; Department of Chemistry and Biochemistry, University of California, Los Angeles, Los Angeles, CA, 90095, USA; UCLA-DOE Institute for Genomics and Proteomics, University of California, Los Angeles, Los Angeles, CA, 90095, USA; Molecular Biology Institute, University of California, Los Angeles, Los Angeles, CA, 90095, USA; Quantitative and Computational Biology Institute, University of California, Los Angeles, Los Angeles, CA, 90095, USA; Eli and Edythe Broad Center of Regenerative Medicine and Stem Cell Research, University of California, Los Angeles, Los Angeles, CA, 90095, USA; Thurston Arthritis Research Center, University of North Carolina, Chapel Hill, NC 27599, USA; Department of Cell Biology and Physiology, University of North Carolina, Chapel Hill, NC 27599, USA; Department of Biostatistics, University of North Carolina at Chapel Hill, Chapel Hill, NC, 27516, USA

## Abstract

Genome-wide association studies (GWAS) have successfully identified 145 genomic regions that contribute to schizophrenia risk, but linkage disequilibrium (LD) makes it challenging to discern causal variants. Computational finemapping prioritized thousands of credible variants, ∼98% of which lie within poorly characterized non-coding regions. To functionally validate their regulatory effects, we performed a massively parallel reporter assay (MPRA) on 5,173 finemapped schizophrenia GWAS variants in primary human neural progenitors (HNPs). We identified 439 variants with allelic regulatory effects (MPRA-positive variants), with 71% of GWAS loci containing at least one MPRA-positive variant. Transcription factor binding had modest predictive power for predicting the allelic activity of MPRA-positive variants, while GWAS association, finemap posterior probability, enhancer overlap, and evolutionary conservation failed to predict MPRA-positive variants. Furthermore, 64% of MPRA-positive variants did not exhibit eQTL signature, suggesting that MPRA could identify yet unexplored variants with regulatory potentials. MPRA-positive variants differed from eQTLs, as they were more frequently located in distal neuronal enhancers. Therefore, we leveraged neuronal 3D chromatin architecture to identify 272 genes that physically interact with MPRA-positive variants. These genes annotated by chromatin interactome displayed higher mutational constraints and regulatory complexity than genes annotated by eQTLs, recapitulating a recent finding that eQTL- and GWAS-detected variants map to genes with different properties. Finally, we propose a model in which allelic activity of multiple variants within a GWAS locus can be aggregated to predict gene expression by taking chromatin contact frequency and accessibility into account. In conclusion, we demonstrate that MPRA can effectively identify functional regulatory variants and delineate previously unknown regulatory principles of schizophrenia.

## Introduction

Schizophrenia is a polygenic neuropsychiatric disorder that affects about 24 million people world-wide (McCutcheon et al., 2020). Heritability estimates of schizophrenia are 60-80%, indicating a strong contribution of genetic variation to risk for the disorder (Sullivan et al., 2003). Common variation explains a significant portion of heritability (24% of SNP heritability), and the most recent genome-wide association study (GWAS) has identified 294 genome-wide significant (GWS) loci (Schizophrenia Working Group of the Psychiatric Genomics Consortium et al., 2020). However, it is challenging to understand the functional consequence of these GWS loci, because 1) most reside in non-coding DNA with unknown functions and 2) each GWS locus contains dozens of variants that show significant association due to linkage disequilibrium (LD).

Therefore, a critical step to bridging the gap between genetic loci and biological underpinning is to identify causal variants and delineate their functional impact. While computational finemapping approaches have been developed to predict putative causal variants (Schaid et al., 2018), these methods merely narrow down the search space of causal variants by modeling their decay with LD rather than functionally validating variants. Moreover, different finemapping algorithms can provide different sets of finemapped variants (Mah and Won, 2020). The general consensus in the field is that causal variants exert their function by altering gene expression. To accurately discern variants with gene regulatory effects, experimental validation is pivotal.

Here, we employed a massively parallel reporter assay (MPRA) to experimentally verify the difference in allelic regulatory activity between protective and risk alleles of 5,173 schizophrenia-associated finemapped variants (Pardiñas et al., 2018) in the context of neurogenesis. MPRA provides a scalable genetic approach to characterize gene regulatory effects of thousands of variants in a single experiment (McAfee et al., 2022; Mulvey et al., 2021; Tewhey et al., 2016). We identified 439 MPRA-positive variants that showed allelic regulatory activity in human neural progenitors (HNPs). Pre-existing strategies to prioritize causal variants (e.g. selecting SNPs with strongest GWAS association signals, SNPs with the highest finemap posterior probability, SNPs located in regulatory elements, and/or SNPs with higher evolutionary constraints) did not accurately identify MPRA-positive variants. Transcription factor (TF) binding motif analysis revealed that TFs involved in Wnt signaling pathways were enriched for MPRA-positive variants. To link MPRA-positive variants to genes, we tried different genomic approaches: expression quantitative trait loci (eQTLs) and chromatin interaction profiles (Hi-C). We found that eQTLs and Hi-C identify distinct sets of genes with different (epi)genomic properties. In particular, the Hi-C based approach identified genes with functional annotation, higher selective constraints, and regulatory complexities, suggesting that chromatin architecture is instrumental in assigning GWAS variants to their cognate genes. Consequently, we propose an accessibility by contact model that supplements chromatin contexts to MPRA-measured allelic activity and demonstrate that this model can effectively translate variant function to targetable gene expression.

## Results

### MPRA on schizophrenia risk variants

Since schizophrenia genetic risk factors are enriched in regulatory elements of the developing cortex (Pratt and Won, 2022; Sey et al., 2020; Spiess and Won, 2020; de la Torre-Ubieta et al., 2018), we conducted MPRA in HNPs that model human neural development (Stein et al., 2014) (**Figure 1A**). To perform MPRA in HNPs, we built an AAV-based MPRA vector (AAV-MPRA) that comprises a 150 base-pair (bp) target sequence with the variant in the center, a minimal promoter, GFP, and a 20 bp unique barcode (**Figure 1A, Methods**).

**Figure 1.**
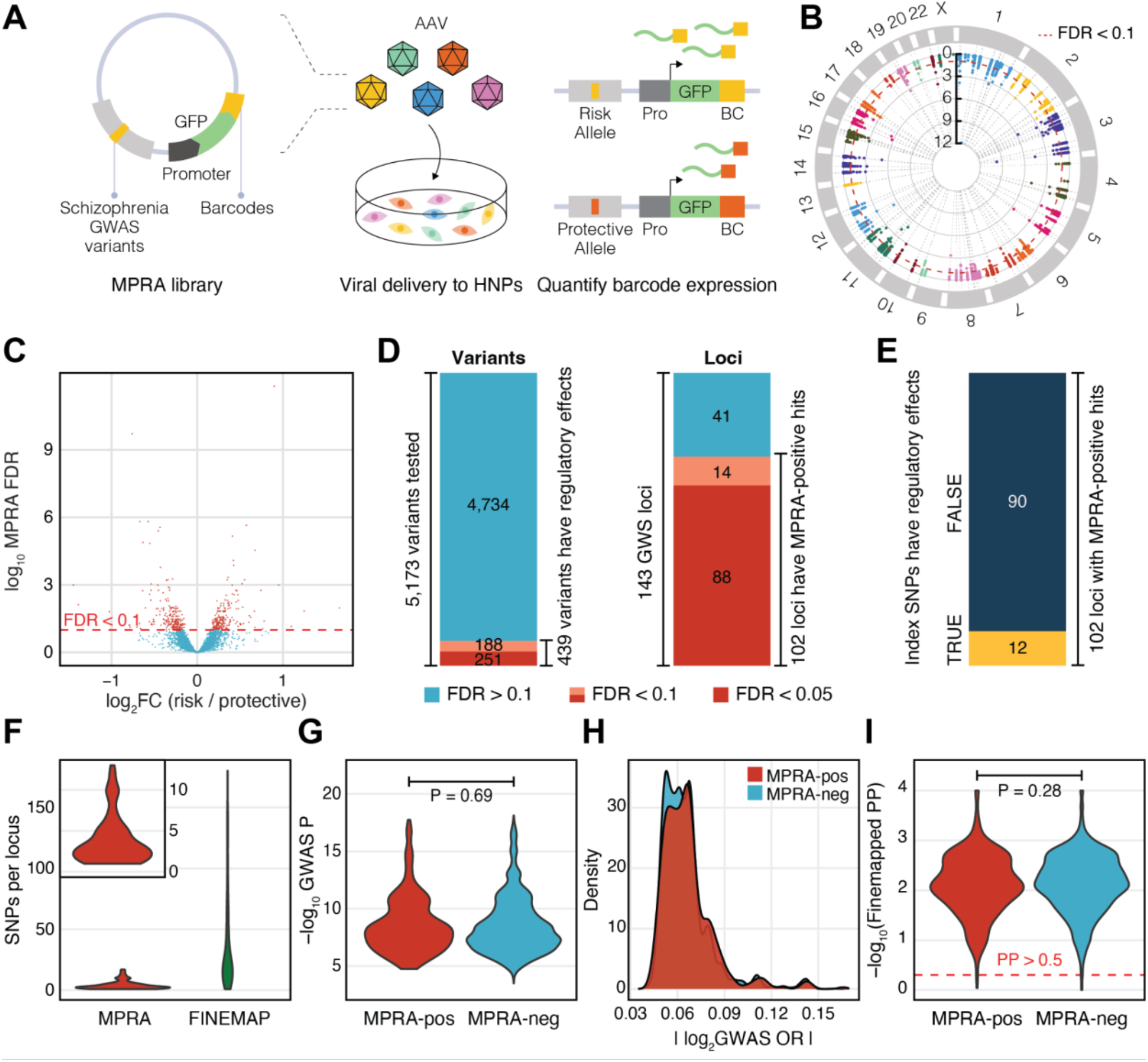
MPRA on schizophrenia risk variants identifies functional regulatory variants. **A**. We generated an MPRA library that contains 5,173 schizophrenia GWAS variants upstream to the promoter, reporter gene, and 20bp barcode. This library was packaged into adeno-associated virus (AAV), which was used to transduce human neural progenitors (HNPs). We compared barcode expression counts between risk and protective alleles to identify variants that show allelic regulatory activity (MPRA-positive variants). **B**. We display our MPRA results within a circular manhattan plot. Red dotted line indicates FDR=0.1. **C**. Volcano plot showing allelic regulatory activity of 5,173 finemapped credible variants. **D**. Out of 5,173 fine-mapped credible variants from 143 genome-wide significant (GWS) loci, 439 variants exhibited allelic regulatory effects in HNPs covering 102 GWS loci (FDR<0.1). **E**. Out of 102 GWS loci with regulatory activity, only 12 GWAS index variants showed allelic regulatory activity. **F**. MPRA dramatically reduced the number of causal variants per locus. **G-I**. GWAS association P-values (**G**), GWAS odds ratio (OR, **H**), and finemap posterior probabilities (**I**) do not differ between MPRA-positive (MPRA-pos) and MPRA-negative (MPRA-neg) variants. P-values were calculated by the Wilcoxon rank sum test.

Using this AAV-MPRA backbone, we generated an AAV-MPRA library for a computationally predicted credible set of schizophrenia risk variants (Pardiñas et al., 2018). We compiled 150 bp target sequences centered on 6,064 finemapped schizophrenia risk variants (**Supplementary Figure 1A-B**). Among them, 470 target sequences that harbor either risk variants larger than 10 bp or recognition sites for restriction enzymes used in the cloning steps were removed (**Supplementary Figure 1A, Methods**). After filtering out low quality and/or undetected variants, 5,173 variants (10,346 risk and protective alleles) were included in the final AAV-MPRA library that covers 143 out of 144 GWS loci (**Supplementary Figure 1A**).

Because the size of variants (<10 bp) is smaller than the barcodes (20 bp), we reasoned that the effects of barcodes on GFP expression can be larger than allelic regulatory effects. To control for the potential effects of barcodes on GFP expression and to fully capture the small effect size of allelic regulatory activity, we mapped each allele to 185 barcodes on average (**Supplementary Figure 2A**).

The resulting schizophrenia MPRA library was packaged into the AAV, which was administered to HNPs. Two weeks after administering the AAV-MPRA library to HNPs, RNA was extracted from the transduced cells and barcoded GFP expression was quantified by RNA sequencing (RNA-seq). RNA barcode counts were aggregated for a given allele to obtain summarized allelic expression. To control for transduction efficiency and barcode dispersion during cloning, DNA barcode counts from the AAV-MPRA library were used for normalization (**Methods**). The correlation coefficients across biological replicates ranged from 0.57 to 0.75 (**Supplementary Figure 2B-C**).

To identify finemapped variants with allelic regulatory activity, RNA barcode counts for protective and risk alleles in a total of 10 biological replicates were compared against the corresponding viral DNA barcode counts using the *mpra* Bioconductor package (**Figure 1B, Methods**). As a result, we identified 439 variants that show allelic regulatory activity at an FDR threshold of 0.1 (hereafter referred to as MPRA-positive variants, **Figure 1C-D, Supplementary Table 1**). We found that 102 out of 143 GWS loci contained at least one MPRA-positive variant (**Figure 1D**). Out of 102 GWS loci that harbor regulatory variants, index variants (variants with the strongest GWAS association statistics at given loci) of 12 loci showed regulatory activity (**Figure 1E**), suggesting that the most significant GWAS association cannot accurately predict functional variants.

MPRA not only refined the number of regulatory variants, but also narrowed down the number of variants per locus (**Figure 1F**). On average, 36.2 variants per locus were identified via computational finemapping approaches. MPRA further pruned them to 4.30 variants per locus. Moreover, 18 out of 102 loci could be pinpointed to a single regulatory variant, demonstrating the power of MPRA in refining GWS loci.

We next evaluated whether association statistics from GWAS or computational finemapping may distinguish MPRA-positive variants from MPRA-negative variants (see **Methods** for their definition). We found that MPRA-positive variants did not differ from MPRA-negative variants in their GWAS association statistics such as P-values and effect sizes (**Figures 1G-H**). Similarly, finemap posterior probabilities did not differ between MPRA-positive and -negative variants (**Figure 1I**). These results show that predictive models purely based on statistical associations do not accurately predict regulatory effects of finemapped variants associated with schizophrenia.

### Epigenetic properties of MPRA-positive variants

To further characterize MPRA-positive variants, we surveyed genomic annotations of MPRA tested variants (**Supplementary Figure 3A**). As expected, the majority of MPRA tested variants were located in intergenic and intronic regions, and only a small portion of them were located in exons and promoters. We did not observe a clear distinction between MPRA-positive and - negative variants in their genomic annotation.

We next sought to characterize epigenetic properties of MPRA-positive and -negative variants. Schizophrenia heritability is enriched in brain and neuronal enhancers (Sey et al., 2020), supporting the prevailing hypothesis in GWAS that causal variants are more likely located in enhancer regions. Therefore, we compared the enhancer overlap between MPRA-positive and - negative variants (**Figure 2A-C**). Notably, MPRA-positive and -negative variants overlapped with adult and fetal brain enhancers (Li et al., 2018) at a similar proportion (**Figure 2A**). Similarly, MPRA-positive and -negative variants did not significantly differ in their overlap with cell-type specific enhancers identified in the postnatal brain (Nott et al., 2019) (**Figure 2B**) and the prenatal brain (Ziffra et al., 2021) (**Figure 2C**). The only exception was early excitatory neuronal enhancers of the prenatal brain, in which MPRA-positive variants were more frequently located compared to MPRA-negative variants (**Figure 2C**).

**Figure 2.**
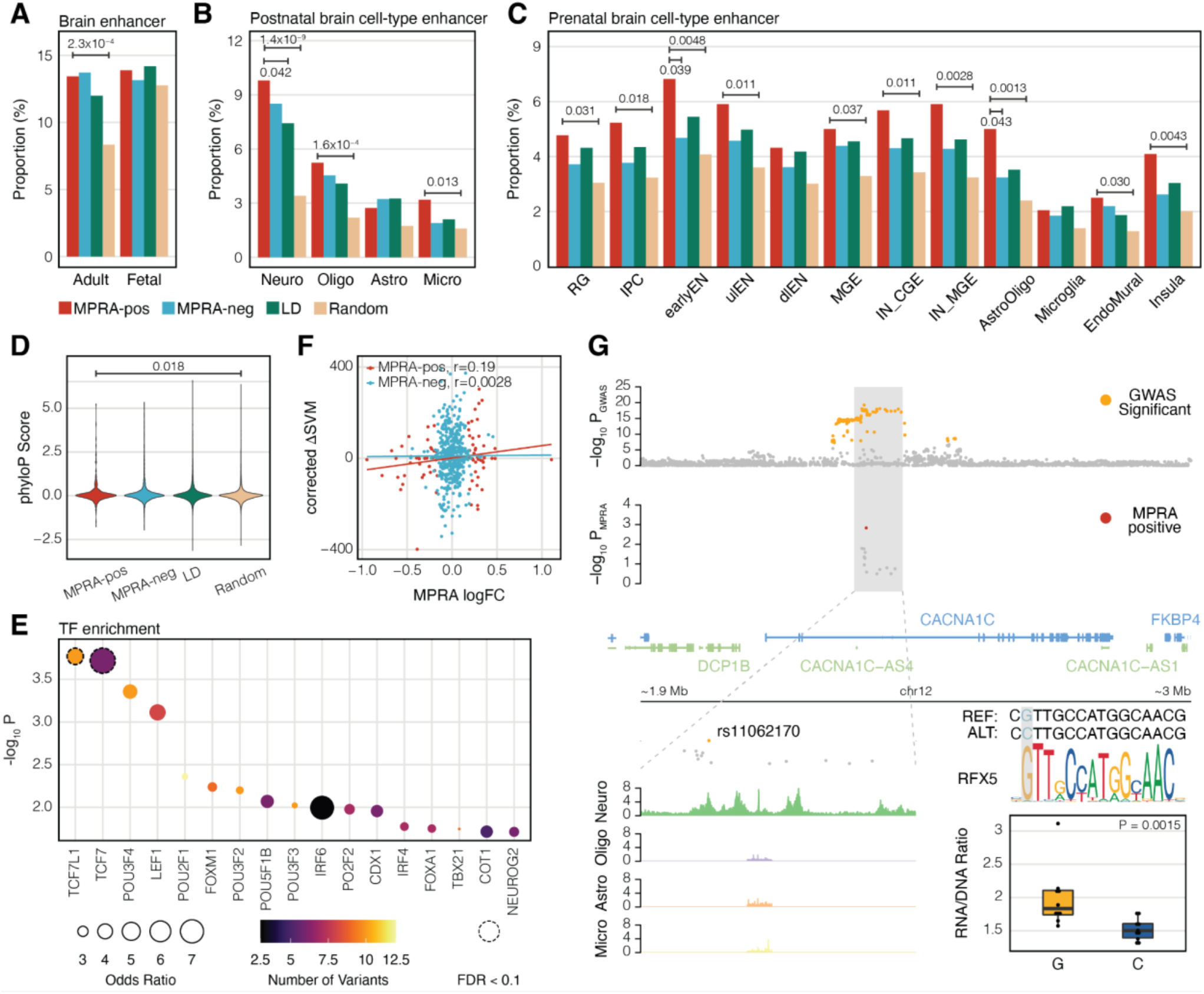
Epigenetic characterization of MPRA-positive schizophrenia risk variants. **A-C**. The proportion of epigenetic overlap of MPRA-positive (MPRA-pos) and MPRA-negative (MPRA-neg) variants, LD SNPs, and random SNPs to the adult and fetal brain enhancers (**A**), cell-type specific enhancers in the adult brain (**B**), and cell-type specific enhancers in the fetal brain (**C**). P-values were calculated by one-sided Fisher’s exact test. Comparisons have been made between MPRA-positive variants and other sets of variants. Only significant P-values are depicted. Neuro, neurons; Oligo, oligodendrocytes; Astro, astrocytes; Micro, microglia; RG, radial glia; IPC, intermediate progenitor cells; earlyEN, early excitatory neurons; ulEN, upper-layer excitatory neurons; dlEN, deep-later excitatory neurons; MGE, medial ganglionic eminence; CGE, caudal ganglionic eminence; IN, inhibitory neurons. **D**. Evolutionary conservation (phyloP scores) of MPRA-positive, MPRA-negative, LD, and random SNPs. **E**. TFs whose motifs are predicted to be altered by MPRA-positive variants. TF enrichment was calculated by comparing TF binding motifs between MPRA-positive variants and LD SNPs. Each dot is color-coded based on the number of variants that are predicted to alter TF binding motifs and the size of the dot represents the odds ratio. Dotted circles represent TFs that meet the FDR threshold (FDR<0.1). **F**. Expression outcome of MPRA (measured by MPRA logFC) can be predicted by the combination of TF binding, activity, and expression (measured by corrected *Δ*SVM) for MPRA-positive variants, but not for MPRA-negative variants. r stands for Pearson’s correlation coefficient. **G**. Integration of MPRA and other functional genomic datasets unveils a causal variant (rs11062170), a trans regulator (RFX5), and a cell type (neuron) for the *CACNA1C* locus. The MPRA-positive SNP rs11062170 lies within H3K27ac peaks of neurons but not other cell types. The alternative allele (C) of rs11062170 breaks the binding motif of RFX5 and is correlated with lower expression of a reporter gene in MPRA.

This is not contradictory to the previous finding that schizophrenia genetic risk factors are enriched in brain and neuronal enhancers, because MPRA-positive variants more frequently overlapped with brain (**Figure 2A**) and neuronal (**Figures 2B-C**) enhancers than the genomic background (random SNPs: SNPs that are matched for minor allele frequency [MAF] and LD). In contrast, MPRA-positive variants did not display elevated epigenetic overlap compared with the local background (LD SNPs: non-finemapped SNPs within schizophrenia GWS loci). MPRA-positive variants also showed cell-type specificity, with the highest level of overlap with neuronal enhancers within the postnatal brain (**Figure 2B**) and early excitatory neuronal enhancers within the prenatal brain (**Figure 2C**) among other cell types. We believe that the observed cell-type specificity is not purely driven by the cell line (i.e. HNPs) in which MPRA was conducted, because progenitors (e.g. radial glia and intermediate progenitor cells that HNPs resemble) would be expected to have higher epigenetic overlap than neurons. Together, these results show that MPRA-positive variants are enriched in neuronal regulatory architecture compared to the genomic background, but epigenetic properties alone cannot predict functional regulatory variants from non-regulatory variants.

It has been previously reported that schizophrenia GWAS signals are under strong selective pressure (Pardiñas et al., 2018). We therefore explored the evolutionary conservation of variant-harboring regions (150 bp regions centered on each variant) for MPRA-positive, MPRA-negative, LD, and random SNPs (**Figure 2D**). We employed evolutionary conservation scores calculated by comparative genomic analyses across 240 species (Zoonomia Consortium, 2020). MPRA-positive variants showed elevated evolutionary constraints compared to random SNPs albeit to a small degree (Wilcoxon rank sum test, P=0.018). On the contrary, evolutionary constraints did not differ between MPRA-positive and -negative variants (Wilcoxon rank sum test, P=0.46) or between MPRA-positive variants and LD SNPs (Wilcoxon rank sum test, P=0.86). Similar to this result, PhastCons scores, another metric for evolutionary conservation, did not differ between MPRA-positive variants and other sets of SNPs (**Supplementary Figure 3B**).

Because we are using an episomal version of MPRA, the allelic regulatory activity is mainly driven by TFs. We used motifbreakR (Coetzee et al., 2015) to identify TFs whose binding motifs are predicted to be disrupted or created by each set of variants (**Supplementary Table 2**). We then identified TFs whose binding motifs are enriched in MPRA-positive variants compared to the local background (**Figure 2E**) or the global background (**Supplementary Figure 3C**). Top TFs enriched for MPRA-positive variants include the T-cell factor/lymphoid enhancer factor (TCF/LEF) family (e.g. TCF7, TCF7L1, and LEF1). TCF7 and LEF1 are major mediators of Wnt signaling (Cadigan and Waterman, 2012). We have previously found that genetic variants near Wnt signaling pathways were associated with various psychiatric disorders including schizophrenia and autism spectrum disorder (ASD) (Sey et al., 2020). Therefore, these results highlight the significance of Wnt signaling pathways in understanding the genetic etiology of psychiatric disorders.

To further address the relationship between TF binding and allelic regulatory activity, we leveraged 94 high-confidence delta support vector machine (*Δ*SVM) frameworks that predict variants with differential binding to TFs (Yan et al., 2021). Because TFs can act as activators or repressors, TF activity needs to be taken into account in translating TF binding to regulatory activity. Moreover, highly expressed TFs can have a larger impact on SNP-mediated regulatory activity than lowly expressed TFs. Consequently, we calculated corrected *Δ*SVM for each variant by combining preferential allelic binding (*Δ*SVM scores), expression levels, and activity (1 if a TF is an activator and –1 if a TF is a repressor) of TFs (see **Methods** for the equation). Corrected *Δ*SVM scores were moderately correlated with allelic regulatory activity (measured by MPRA logFC) for MPRA-positive variants, but not for MPRA-negative variants (**Figure 2F, Supplementary Figure 3D**). This result suggests that TFs are key drivers of allelic regulatory activity measured by MPRA. Given that *Δ*SVM frameworks have been established for only 94 TFs, we expect that the allelic regulatory activity could be better modeled when we have a more complete understanding of TF-SNP interaction.

In an example of tying together MPRA results and epigenetic profiles, we highlight a MPRA-positive variant, rs11062170, in the *CACNA1C* locus (**Figure 2G**). This variant is located within a H3K27ac peak for neurons (Nott et al., 2019), but not other brain cell types, alluding to the variant’s neuronal specificity. This variant is located within the intron of *CACNA1C*, a gene that encodes a voltage-gated calcium channel. *CACNA1C* was previously identified to be associated with schizophrenia (Roussos et al., 2014). Allelic regulatory activity of rs11062170 measured by MPRA showed that the reference (protective) allele, G, induced significantly higher expression than the alternative (risk) allele, C. The alternative allele is predicted to break the binding motif of RFX5, alluding to the mechanism of action of lower expression for the alternative allele.

### Cell-type specificity of MPRA results

The observed cell-type specificity of MPRA-positive variants (**Figure 2B-C**) encouraged us to compare our results with previously published MPRA data obtained from K562 lymphoblast and SY5Y neuroblastoma cell lines (Myint et al., 2020). K562 lymphoblasts are a non-neuronal cell line, and SY5Y neuroblastoma were previously reported to display transcriptomic profiles that poorly resemble *in vivo* brain development as compared to HNPs as used here (Stein et al., 2014). We therefore hypothesized that MPRA-positive variants identified from HNPs will be distinct from MPRA-positive variants from other cell lines. Out of 5,173 variants tested in our MPRA, only 565 variants were tested in K562 and SY5Y due to the difference in SNP selection strategies (**Supplementary Figure 4A**). We detected 49, 40, 104 variants to have allelic regulatory activity in HNPs, SY5Y, and K562, respectively (FDR<0.1 using 565 variants tested in both studies, **Supplementary Figure 4A**). A minimal overlap of MPRA-positive variants was detected when comparing HNPs with SY5Y (4 variants, **Supplementary Figure 4B**) and HNPs with K562 (11 variants, **Supplementary Figure 4C**). While we cannot rule out other contributing factors (e.g. batch effects, different experimental strategies, different statistical analysis), this result suggests that variant effects on gene regulation may substantially differ by cell types.

### MPRA identifies a different set of variants from eQTLs

eQTLs have become the primary genomic resource to functionally link GWAS to gene expression measures (GTEx Consortium, 2020). Since MPRA identifies allelic regulatory activity of variants as eQTLs do, we compared MPRA-positive variants with eQTLs detected in the adult prefrontal cortex (PFC) (Wang et al., 2018). Notably, only 36% of MPRA-positive variants showed eQTL signals (**Figure 3A**). Among 157 variants with both MPRA allelic regulatory activity and eQTL signals, 130 variants (83%) exhibited the identical direction of the effect (hereby referred to as IDE variants, **Supplementary Table 3**), indicating a high level of concordance between MPRA and eQTLs when both signals are detected. Because HNPs better model developing brains than adult brains, we also compared MPRA-positive variants with eQTLs from the developing brain (Walker et al., 2019). Comparison with developing brain eQTLs gave similar findings, albeit to a lesser degree of overlap, which could be due to the low detection power of developing brain eQTLs from lower sample size and other factors (**Supplementary Figure 5**). Because adult brain eQTLs (238,194 eQTLs associated with 32,944 genes) are better powered than developing brain eQTLs (7,962 eQTLs associated with 6,526 genes), we used adult brain eQTLs for the rest of the analysis.

**Figure 3.**
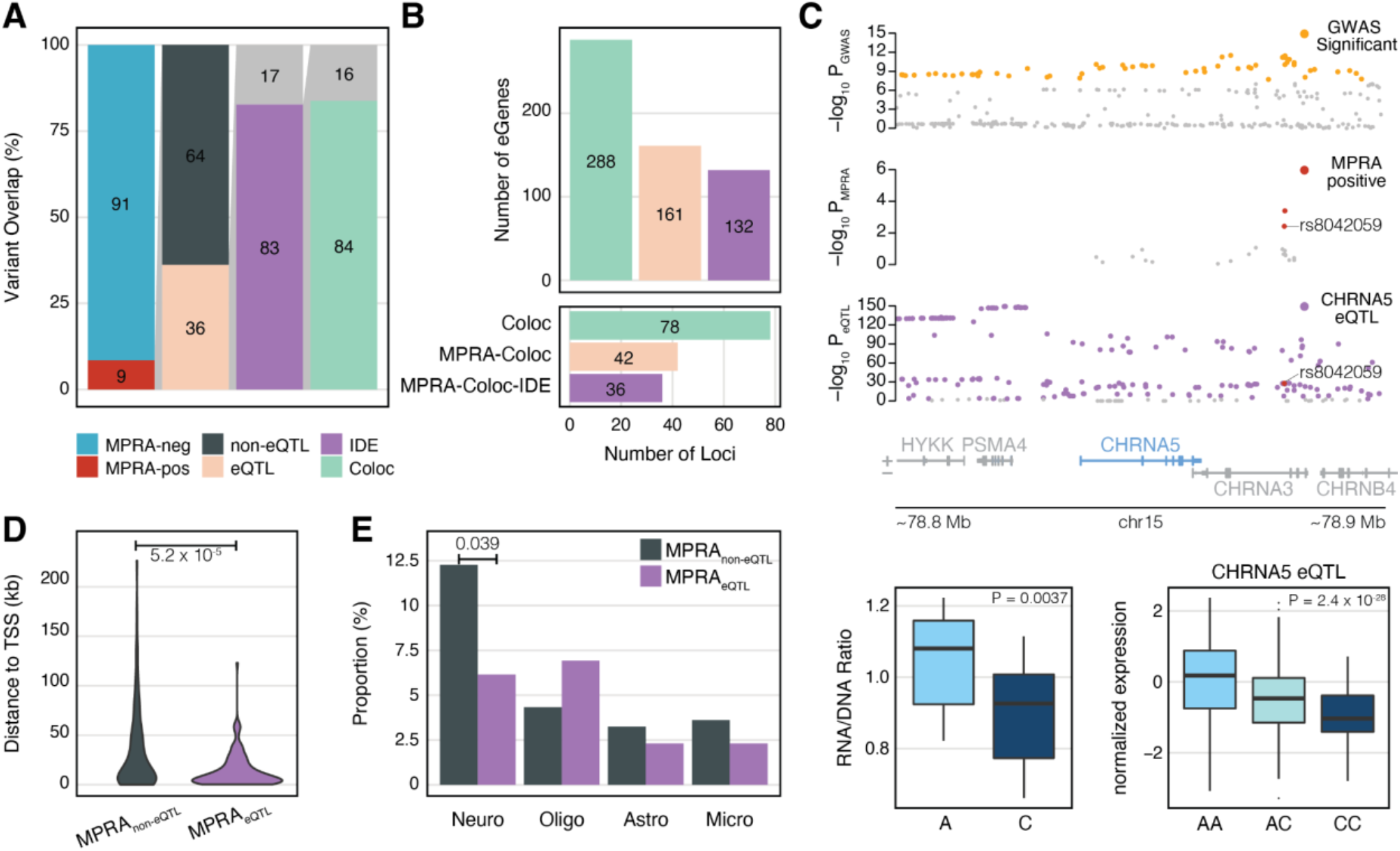
Comparison of MPRA results with adult brain eQTLs. **A**. 9% of the variants tested in our MPRA were found to have allelic regulatory effect (MPRA-positive variants). 36% of MPRA-positive variants overlapped with eQTLs. Within the 36% overlap with eQTLs, 83% of MPRA-positive variants are identical in expression direction (IDE) to the overlapping eQTL variants. Within that 83%, 84% of IDE variant-gene pairs were detected from the colocalization analysis between eQTLs and schizophrenia GWAS (Coloc). **B**. 78 schizophrenia GWS loci colocalize with eQTLs, providing 288 schizophrenia-associated eGenes (Coloc). 42 out of these 78 loci contain at least one MPRA-positive variant and are mapped to 161 eGenes (MPRA-Coloc). 36 of MPRA-Coloc loci contain variants that have the identical direction of effects between MPRA and eQTLs and are mapped to 132 eGenes (MPRA-Coloc-IDE). **C**. eQTLs for the *CHRNA5* gene colocalize with a schizophrenia GWS locus on chromosome 15. Within this locus are two MPRA-positive variants. One of the MPRA-positive variants, rs8042059, shows the identical direction of effects between MPRA and eQTLs that the alternative allele C is associated with downregulation of *CHRNA5*. **D**. MPRA variants that do not overlap with eQTL (MPRA_non-eQTL_) and MPRA variants that do overlap with eQTL (MPRA_eQTL_) differ in distance to the transcription start site (TSS). **E**. MPRA_eQTL_ variants more frequently overlap with neuronal enhancers compared to MPRA_non-eQTL_ variants. P-values were calculated by one-sided Fisher’s exact test.

Because eQTLs are affected by LD, simple genomic coordinate-level overlap between GWAS and eQTLs could lead to spurious overlap. Colocalization analysis has been implemented to evaluate whether GWAS and eQTLs are explained by a shared set of variants. To test how many IDE variants are also identified from the colocalization analysis, we compared IDE variants with colocalization between schizophrenia GWAS and adult brain eQTLs (Liu et al., 2021) using the *coloc* package (**Methods**). Because colocalization does not always indicate a specific variant, we tested how often eGenes (genes detected as having an associated eQTL) linked to IDE variants were also observed from colocalization analysis. From this analysis, 84% of IDE variants were linked to the same genes as predicted by colocalization analysis (**Figure 3A**).

Intersection of MPRA and eQTLs also pruned the gene list (**Figure 3B**). We initially detected 288 eGenes to be associated with schizophrenia by colocalization analysis, covering 78 loci (Liu et al., 2021). An orthogonal analysis of coordinate-level overlap between eQTLs and MPRA identified 269 eGenes, covering 80 loci. We found that 161 eGenes were shared between colocalization and MPRA-eQTL overlap analysis. After pruning them further with the identical direction of effects between MPRA and eQTLs, 132 eGenes were detected, covering 36 loci.

In an example of MPRA-eQTL IDE overlap, we highlight a schizophrenia GWS locus on chromosome 15 (**Figure 3C**). Two variants at this locus – rs11418931 and rs8042059 – had significant MPRA allelic activity. Rs11418931 was missing in the eQTL analysis, while rs8042059 was detected as an eQTL for a nearby gene, *CHRNA5*. When comparing the directionality of the allelic expression of rs8042059, the reference (risk) A allele increased expression in comparison to the alternate (protective) C allele both in MPRA and eQTL for *CHRNA5*.

Mostafavi et al. have recently postulated that eQTL studies and GWAS identify a different set of variants (Mostafavi et al., 2022). In their analysis, variants detected in eQTL studies and GWAS differ by their distance to transcription start sites (TSS) and regulatory architecture. To investigate whether MPRA could identify a distinct set of disease-associated variants that are not explained by variants detected as eQTL, we characterized genomic and epigenomic properties of MPRA-positive variants with and without eQTL signature (hereafter referred to as MPRA_eQTL_ variants and MPRA_non-eQTL_ variants, respectively, see **Methods** for how they were defined).

When comparing the distance of MPRA_eQTL_ and MPRA_non-eQTL_ variants to the TSS, we found that MPRA_non-eQTL_ are more distal to the TSS than MPRA_eQTL_ variants, hinting that these variants could be involved in distal regulatory relationship (**Figure 3D**). Because distal regulatory elements often encode enhancers, we next surveyed whether there is a difference between MPRA_eQTL_ and MPRA_non-eQTL_ variants in their enhancer overlap. We found that a higher proportion of MPRA_non-eQTL_ variants (∼12%) overlapped with neuronal enhancers compared to MPRA_eQTL_ variants (∼6%) (Nott et al., 2019). Such difference in enhancer overlap was not shown in other tested cell types (**Figure 3E**). Taken together, these results suggest that disease-associated variants may differ from variants detected as eQTL, and MPRA could fill this gap by testing GWAS variant effects on gene regulation in a manner independent of issues related to eQTL study power.

### Identification of schizophrenia candidate risk genes via long-range chromatin interactions

Since MPRA-positive variants exhibited different epigenomic properties from eQTLs (**Figure 3D-E**), we sought another method for assigning target genes for MPRA-positive variants. The previous finding that genes affected by GWAS variants show enhanced regulatory complexity (Mostafavi et al., 2022) prompted us to leverage long-range chromatin interaction datasets from the human brain. As MPRA-positive variants were preferentially located in enhancers of immature and mature neurons (**Figure 2B-C, Figure 3E**), we used chromatin loops in neural progenitors (germinal zone, GZ), immature postmitotic neurons (cortical plate, CP), pediatric neurons, and adult neurons (**Figure 4A**) (Hu et al., 2021; Nott et al., 2019; Won et al., 2016). Neuronal chromatin interaction datasets assigned 209 MPRA-positive variants to 272 protein-coding genes (hereby referred to as MPRA_Hi-C_ genes, **Supplementary Table 4**). The resulting SNP-gene relationship was multivalent. On average, each SNP was mapped to 2.7 genes (**Figure 4B**), while each gene was mapped to 2.1 MPRA-positive SNPs (**Figure 4C**). Only 24 genes overlapped between the MPRA_Hi-C_ genes and the MPRA_eQTL-IDE_ genes (**Figure 4D**), showing that the two datasets assign the MPRA-positive variants to distinct sets of genes.

**Figure 4.**
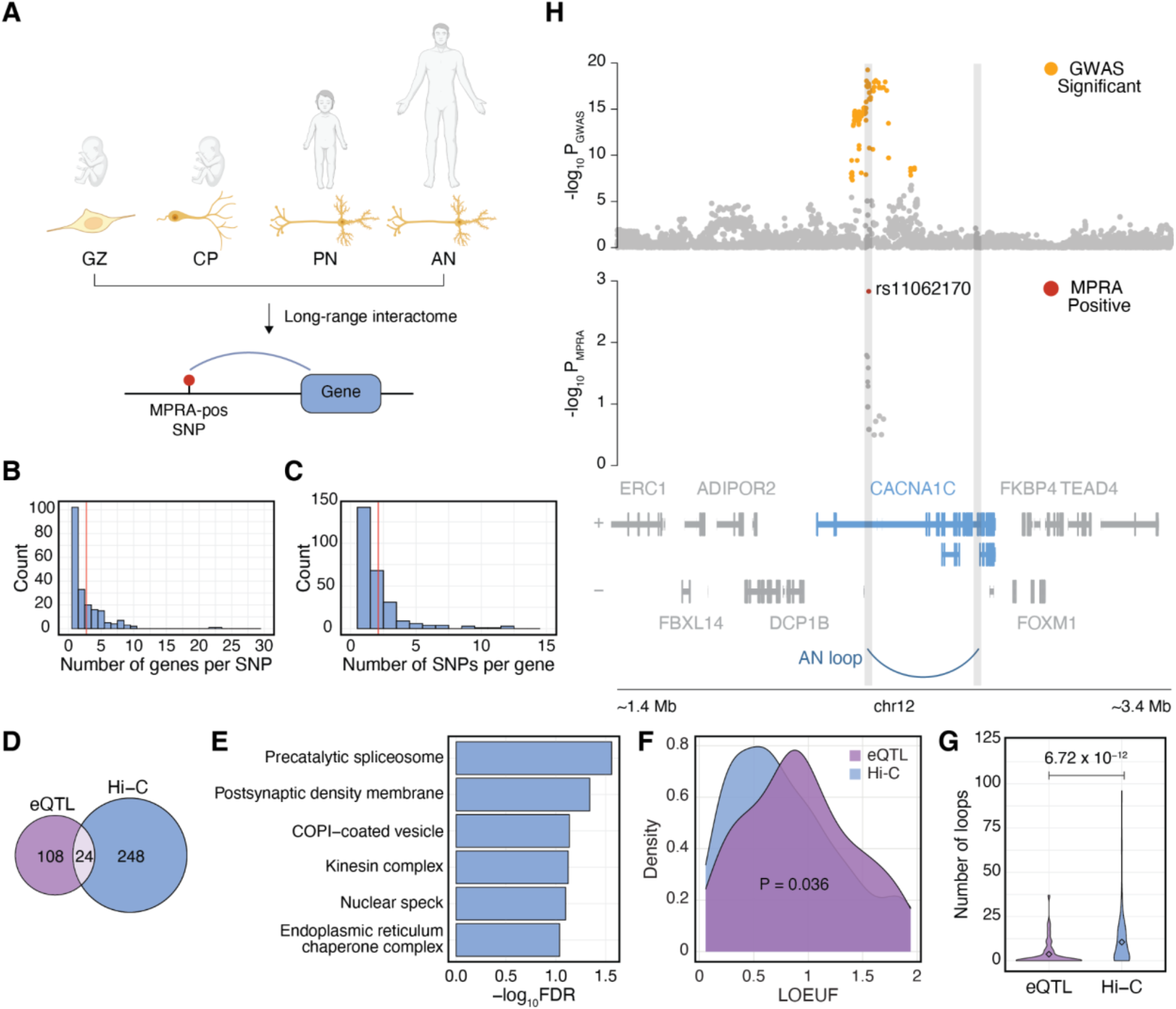
Genes assigned to MPRA-positive variants using long-range interactome differ from eQTL-assigned genes. **A**. Chromatin loops from neurons of four different developmental time points were used to map MPRA-positive variants to genes. GZ, germinal zone; CP, cortical plate; PN, pediatric neuron; AN, adult neuron. **B**. Distribution of the number of genes mapped per SNP. Red line, mean. **C**. Distribution of the number of variants mapped per gene. Red line, mean. **D**. Overlap between MPRA_eQTL-IDE_ genes and MPRA_Hi-C_ genes. **E**. Gene ontology (GO) analysis of MPRA_Hi-C_ genes indicates involvement of spliceosome and synaptic functions in schizophrenia etiology. Redundant GO terms were omitted (see **Supplementary Figure 6A** for full GO terms). **F**. Loss-of-function observed/expected upper bound fraction (LOEUF) score distribution shows that MPRA_Hi-C_ genes are less tolerant to mutations compared to MPRA_eQTL-IDE_ genes. **G**. The number of promoter-anchored loops show higher regulatory complexity for MPRA_Hi-C_ genes compared to MPRA_eQTL-IDE_ genes. Loops from adult neurons were used. P-values calculated by Wilcoxon rank sum test. **H**. An example locus for *CACNA1C* shows that the MPRA-positive SNP rs11062170 physically interacts with *CACNA1C* promoter in adult neurons.

MPRA_Hi-C_ genes were enriched for gene ontologies (GO) related to spliceosomes and synaptic functions (**Figure 4E, Supplementary Figure 6A**). Enrichment of MPRA_Hi-C_ genes in spliceosomes corroborates pervasive isoform-level dysregulation in schizophrenia brains (Gandal et al., 2018). Furthermore, synaptic involvement of MPRA_Hi-C_ genes recapitulates a widely accepted notion that neurons are the central cell type for schizophrenia (Sey et al., 2020; Skene et al., 2018). Accordingly, MPRA_Hi-C_ genes showed elevated expression in neurons than non-neuronal cells both in the fetal (**Supplementary Figure 6D**) and adult cortex (**Supplementary Figure 6E**).

One of the genes that physically interacts with MPRA-positive variants was *CACNA1C* (**Figure 4H)**. Chromatin interaction offers a complete mechanism of action for the *CACNA1C* locus (also depicted in **Figure 2G**): The MPRA-positive SNP rs11062170, located within a neuronal enhancer (**Figure 2G**), interacts with the promoter of *CACNA1C* in adult neurons (**Figure 4H**). The alternative (risk) allele C of rs11062170 disrupts RFX5 binding, which weakens the neuronal enhancer activity (**Figure 2G**). The weakened neuronal enhancer propagates to the decreased expression of *CACNA1C* via a neuronal chromatin loop.

Another example is *KCNG2* locus (**Supplementary Figure 7**). In this locus, MPRA-positive SNP rs11664298 interacts with the promoter of *KCNG2* through a chromatin loop from neural progenitors. The alternative allele A of rs11664298 is associated with significant downregulation of the reporter gene compared with the reference allele G. Furthermore, the alternative allele is also a risk allele for schizophrenia, suggesting downregulation of *KCNG2* in schizophrenia. In line with this, *KCNG2* was downregulated in postmortem brains of schizophrenia-affected individuals (Gandal et al., 2018). The alternative allele A of rs11664298 breaks a ZNF121 binding motif, providing a potential trans-regulatory mechanism of action for this locus. Collectively, we found that 38% of MPRA-positive variants (167/439) interact with promoters through chromatin loops and also alter TF binding motifs (**Supplementary Table 5**), suggesting long-range interaction as a mechanism for how variants alter gene expression.

Mostafavi et al. have shown that genes linked to variants detected in eQTL studies and GWAS differ by their functional annotation, mutational constraint, and regulatory complexity (Mostafavi et al., 2022). In their study, all eGenes, regardless of disease association, were compared against genes proximal to GWAS variants, so it is unclear whether genes linked to GWAS variants also differ when different mapping strategies were used. Given the epigenetic difference between MPRA_eQTL_ variants and MPRA_non-eQTL_ variants, we hypothesized that genes assigned to MPRA-positive variants via chromatin interactions (272 MPRA_Hi-C_ genes, **Figure 4D**) differ from those assigned by eQTLs (132 MPRA_eQTL-IDE_ genes, **Figure 3A, 4D**).

Unlike MPRA_Hi-C_ genes that showed functional annotations related to synaptic biology (**Figure 4E, Supplementary Figure 6A**), MPRA_eQTL-IDE_ genes were enriched for more generic cellular function (**Supplementary Figure 6B**). Furthermore, MPRA_Hi-C_ genes exhibited higher mutational constraints than MPRA_eQTL-IDE_ genes (**Figure 4F**), which is in line with the previous report that schizophrenia-associated common variation is enriched for mutation intolerant genes (Pardiñas et al., 2018). Finally, MPRA_Hi-C_ genes were engaged in more distal interaction than MPRA_eQTL-IDE_ genes (**Figure 4G, Supplementary Figure 6C**), indicative of higher regulatory complexity. Taken together, these results suggest that gene assignment for GWAS variants may require an additional annotation strategy utilizing physical interactome, given the significant differences in properties of genes assigned by two different strategies (eQTL vs Hi-C).

### Regulatory principles of multi-variant loci

Out of 102 GWAS loci with functional regulatory variants, only 18 loci were mapped to a single functional regulatory variant while 84 loci had more than one MPRA-positive variant. We explored regulatory relationships of multi-variant loci by mapping them to target genes with neuronal chromatin interactions (**Figure 4A**). Fifty-eight out of 84 multi-variant loci were mapped to genes, and 49 of them were mapped to more than one gene, indicating potential cases of pleiotropy. Adding to another layer of complexity, multiple variants often converged on a single gene. For example, 22 multi-variant loci converged on a single gene. Together, these results suggest that multi-variant loci are often engaged in a complex regulatory relationship that involves pleiotropy and convergence.

Multi-variant loci pose a challenge in translating variant effects to gene expression. Together, 256 out of 272 MPRA_Hi-C_ genes were selected as putative targets of multi-variant loci. Using these genes, we sought to identify how variant effects can be aggregated to predict changes in gene expression. Because cell-type specific transcriptomic signatures of schizophrenia postmortem brains are not available yet, we used gene expression profiles from schizophrenic brain homogenates as a benchmark (Gandal et al., 2018). Out of 256 putative targets of multi-variant loci, 192 genes showed detectable levels of expression in postmortem brains and were used for comparison between MPRA and schizophrenia postmortem expression. To compare MPRA results with schizophrenia postmortem brain expression profiles, we recalibrated MPRA logFC values (alternative/reference allele) to reflect disease risk (risk allele/protective allele). Consequently, variants with positive logFC(risk/protective) values will increase gene expression, while those with negative logFC(risk/protective) values will decrease gene expression in schizophrenia.

To predict gene expression in multi-variant loci, we first explored a simple additive model in which allelic activity of variants (logFC(risk/protective) values) is added to predict gene expression (**Figure 5A-B**). Out of 192 genes compared between MPRA results and postmortem expression profiles, 107 genes (55.7%, permutation P=0.03) showed the consistent direction of effects. Because variants with higher contact frequency may have larger impacts on gene expression, we next weighted allelic activity by chromatin contact frequency (hereby referred to as a contact model, **Figure 5A**). The number of genes with the consistent direction of effects grew from 107 to 109 (56.8%, permutation P=0.0081) by the use of the contact model (**Figure 5B**). We next reasoned that variants within chromatin accessible regions may have a larger impact on gene regulation. Because our episomal design measures allelic activity without taking chromatinization into account, we weighted allelic activity by chromatin accessibility and contact frequency (hereby referred to as an accessibility by contact model, **Figure 5A**). With this model, 116 genes (60.4%, permutation P=3×10^−4^) showed the consistent direction of effects with postmortem expression (**Figure 5B**). Applying the same model to MPRA-negative variants yielded 105 genes (54.7%, permutation P=0.14) to be in the consistent direction (**Figure 5B**).

**Figure 5.**
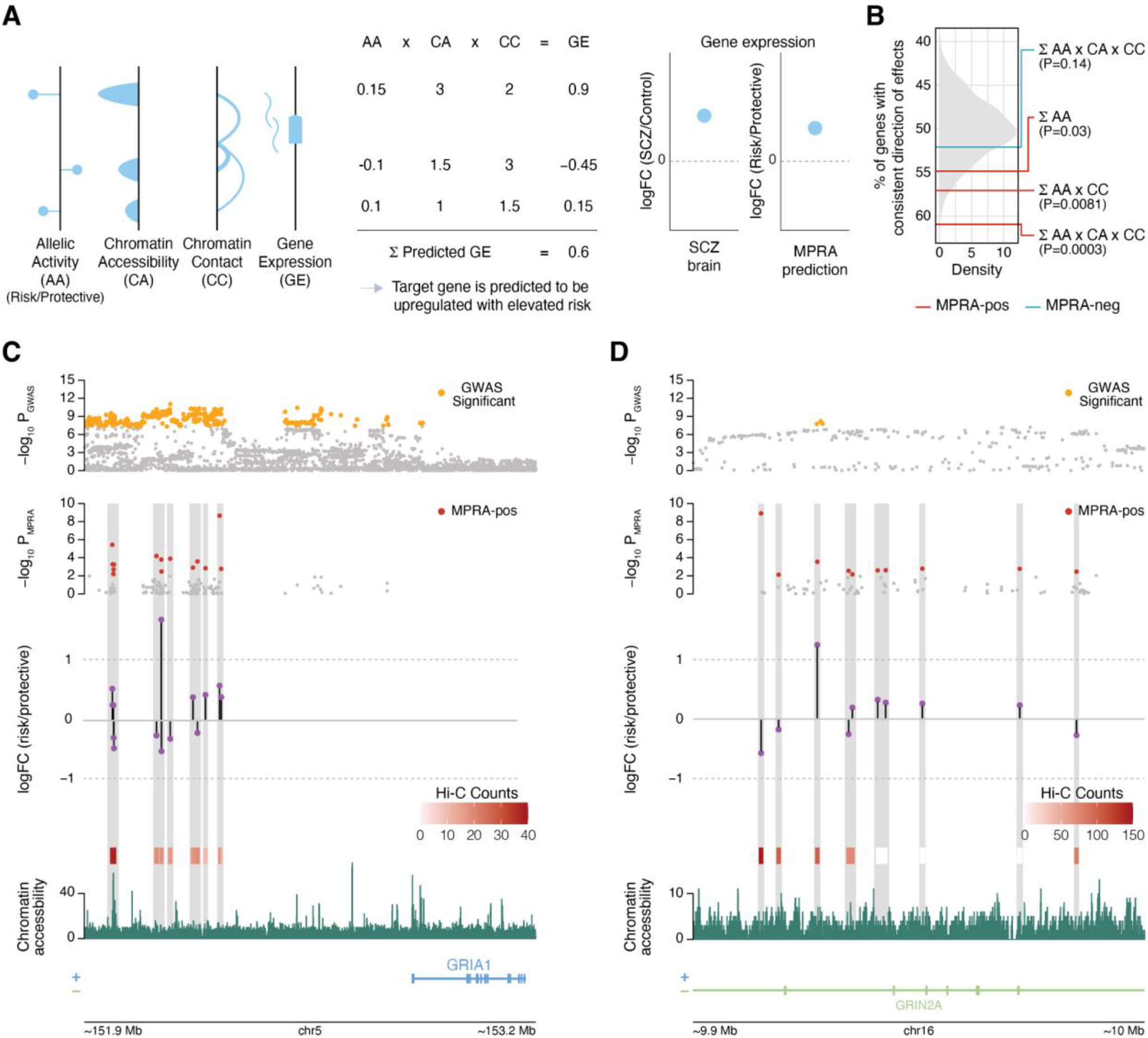
Mapping multi-variant loci to genes. **A**. Illustration of how the accessibility by contact model predicts gene expression outcome for multi-variant loci. **B**. The accessibility by contact model (*∑*allelic activity×chromatin accessibility×chromatin contact) outperforms additive (*∑*allelic activity) and contact (*∑*allelic activity×chromatin contact) models in predicting gene expression changes in schizophrenia postmortem brains. P-values calculated by permutation. **C-D**. *GRIA1* (**C**) and *GRIN2A* (**D**) loci are multi-variant loci in which 14 and 10 variants are predicted to act together on *GRIA1* and *GRIN2A* regulation, respectively. Chromatin contact frequency (Hi-C counts) and accessibility are used to weight MPRA-measured allelic activity (logFC(risk/protective)).

*GRIA1* locus is one of the multi-variant loci in which 14 MPRA-positive SNPs with varying allelic activity may act in concert to regulate *GRIA1* (**Figure 5C**). Both additive and accessibility by contact models predicted that *GRIA1* is upregulated (*∑*logFC=0.25, *∑*logFC×contact×accessibility=0.19), which is consistent with slight upregulation of *GRIA1* in schizophrenia postmortem brains (logFC=0.024). On the other hand, *GRIN2A* locus is an example in which additive and accessibility by contact models give opposite predictions (**Figure 5D**). In this locus, 6 MPRA-positive variants showed detectable levels of chromatin interactions with the *GRIN2A* promoter and included in the model. An additive model suggested that the gene is upregulated (*∑*logFC=1.27), while an accessibility by contact model predicted that the gene is downregulated (*∑*logFC×contact×accessibility=–2.64). *GRIN2A* was modestly downregulated in schizophrenia postmortem brains (logFC=–0.036). Further supporting its downregulation in schizophrenia, *GRIN2A* was shown to have an excess of rare protein-truncating and pathogenic missense variants in schizophrenia (Singh et al., 2022).

In conclusion, combining MPRA allelic activity with chromatin accessibility and contact frequency offers a framework to predict gene expression from MPRA-validated variant effects.

## Discussion

MPRA has demonstrated its ability to vastly narrow down GWAS variants to a list of functionally validated variants with differential allelic activity. We found 439 schizophrenia-associated variants with allelic regulatory effects within 102 GWS loci. Notably, MPRA-positive variants could not be distinguished from MPRA-negative variants by GWAS effect sizes, finemapping posterior probability, evolutionary conservation, or epigenomic annotation such as enhancers. This result highlights the importance of experimental validation in addressing variant function.

The finding that MPRA-positive variants were not necessarily located in enhancers could be due to a number of factors. First, our definition of an enhancer could be incomplete due to the small sample size or shallow read depth. In line with this idea, enhancers defined by scATAC-seq showed a smaller number of overlaps with MPRA-positive variants than enhancers from the bulk brain homogenate. Therefore, MPRA-positive variants may be located in weak enhancers that are yet to be identified. Second, we used an episomal version of MPRA that lacks epigenetic context (e.g. chromatinization). While enhancer activity is heavily dependent on chromatin accessibility, the episomal context of our MPRA enables characterization of variant function without an additional layer of chromatinization. One interpretation is that MPRA exposes allelic activity of variants that do not have relevant *in vivo* function. On the other hand, MPRA can be more sensitive in identifying variants with regulatory activity that may be masked by closed chromatin in the baseline condition. These variants may only be functional under specific regulatory contexts (e.g. upon neuronal activity or cellular stress). While context-specific regulatory variants are difficult to detect via molecular assays in baseline conditions, their implications in disease association are emerging (Alasoo et al., 2018; Umans et al., 2021). Given that the MPRA-measured allelic regulatory effect of a variant depends more so on its binding to TFs than its local chromatin context, we reason that MPRA’s episomal nature may allow us to identify variant priming effects without the need of the stimuli to make the variant accessible.

We looked into TF binding on MPRA-positive variants to understand the mechanism of action of allelic regulatory activity. We found that many differentially expressing alleles either created or broke TF binding motifs and SNP-mediated alterations of TF binding showed moderate predictive values, alluding to the mechanism of action of variant regulatory effects. TFs associated with Wnt signaling were especially interesting as it encodes a major pathway of brain development. Growing evidence suggests the involvement of Wnt signaling in various psychiatric and neurodevelopmental disorders. For example, common variation associated with schizophrenia, depression, and ASD showed enrichment for Wnt signaling pathways (Sey et al., 2020). Altered Wnt signaling was observed in HNPs derived from schizophrenic individuals (Topol et al., 2015).

The pervasive standard in linking variants to gene expression is to leverage eQTL resources. However, a recent paper from Mostafavi et al. suggested that variants detected in eQTL studies may capture a different set of variants than GWAS due to natural selection (Mostafavi et al., 2022). In agreement with this, we found that 64% of MPRA-positive variants did not overlap with variants identified in adult brain eQTL studies. Of the 36% that did overlap, most of them agreed in direction of effects and colocalized with GWAS. It is possible that the little overlap with eQTLs could be due to the differing cell type and developmental stage between eQTLs (heterogeneous adult brain homogenate) and MPRA (HNPs that model neural development), or due to limited sample size of current eQTL studies. Well powered cell-type specific eQTLs (especially neuron-specific eQTLs) may be critical to filling this gap. In contrast, eQTLs from developing brains did not massively differ in their overlap with MPRA-positive variants compared to eQTLs from adult brains.

Despite the potential source of difference, we found that MPRA_non-eQTL_ variants showed different epigenetic properties than MPRA_eQTL_ variants. In particular, MPRA_non-eQTL_ variants were more likely located in distal neuronal enhancers compared with MPRA_eQTL_ variants. This prompted us to employ neuronal distal regulatory relationships (e.g. Hi-C interactions) to link MPRA-positive variants to their cognate genes. MPRA_Hi-C_ genes exhibited richer functional annotation and stronger selective constraints than MPRA_eQTL-IDE_ genes. Moreover, MPRA_Hi-C_ genes were engaged in more distal regulatory interactions, which aligns with the reported enhancer redundancy of disease-associated mutation intolerant genes (Wang and Goldstein, 2020). Collectively, our results suggest that unbiased characterization of GWAS variants via MPRA could identify functional regulatory variants under selective pressure that eQTLs may not be able to detect.

As chromatin architecture provides a complementary approach to map GWAS variants not cataloged by eQTLs, we sought to explore how chromatin architecture can be integrated with allelic activity to predict gene expression from variant regulatory effects within multi-variant loci. We found that the accessibility by contact model outperformed a simple additive model in predicting the direction of gene expression change. This model adds to the recently proposed activity-by-contact model that predicts the relationship between regulatory elements and genes (Fulco et al., 2019). Current prediction accuracy of the accessibility by contact model was ∼60% when compared against the gene expression profile from schizophrenia brain homogenates. Because neuronal chromatin accessibility (Fullard et al., 2018) and contact maps (Hu et al., 2021) were used to translate the functional impact of MPRA-positive variants that are enriched in neuronal enhancers, we expect that the prediction accuracy could be further improved by the comparison with neuronal-specific transcriptomic signatures in schizophrenia. In conclusion, the combination of MPRA-measured allelic activity with chromatin architecture can complement the episomal design of MPRA that does not account for the endogenous genomic context and provide a systematic framework to interpret variant effects on gene regulation.

## Methods

### Variant selection

FINEMAP (Benner et al., 2016) was applied to 144 schizophrenia GWS loci (excluding the MHC locus) from Pardinas et al. (Pardiñas et al., 2018). A set of finemapped variants that can explain a given GWS loci with 95% probability for containing causal configuration was selected as previously described (Schork et al., 2019). In total, we identified 6,064 finemapped variants for 144 schizophrenia GWS loci. A 150bp sequence flanking each variant was then selected to be inserted to the MPRA library. For indels, we used the same sized fragment (150bp) centered to the variant. We found that 150bp flanking sequences of 470 variants out of 6,064 finemapped variants contained sequences for restriction enzymes (MluI, SpeI, KpnI, XbaI) used for molecular cloning. These variants were excluded, resulting in 5,594 variants that were tested via our MPRA framework.

### Creating variant oligo library

The 202 bp library oligos that contain schizophrenia risk variants were synthesized by Agilent and amplified using NEBNext 2X Q5 Hifi HS Mastermix (NEB, cat#M0453S; primers: *MPRA-chipprimer-R* and *MPRA-chipprimer-F*). Primer information is available in **Supplementary Table 6**.

We then used a pair of primers, one with the 20bp random barcode and SpeI restriction site (*MPRA-BC_Primer_R*) and the other with the MluI restriction site (*MPRA-BC_Primer_F*) to add random barcodes and restriction sites to the library oligos via PCR (NEBNext 2X Q5 Hifi HS Mastermix). The resulting library was digested with SpeI-HF (NEB, cat#R3133S) and MluI-HF (NEB, cat#R3198S) for 1 hour at 37°C, followed by rSAP treatment (NEB, cat#M0371S) for 1 hour at 37°C and heat inactivation for 5 minutes at 65°C. After digestion, the library was cleaned up using Zymo clean and concentrator kit (Zymo, cat#D4033).

### Engineering of AAV-MPRA backbone

We obtained the AAV backbone plasmid (pAAV-hSyn-EGFP) from the UNC vector core (https://www.addgene.org/50465/). We digested pAAV-hSyn-EGFP using MluI-HF and EcoR1-HF (NEB, cat#R3101S), and ligated in an oligo that contains the sequences for MluI, SpeI, and EcoRI restriction sites using T7 DNA ligase (NEB, cat#M0318S). The ligated plasmid was transformed into Endura electrocompetent cells (Lucigen, cat#60242-1) via electroporation and grown in ampicillin LB overnight at 30°C. The cells were mini prepped with Qiagen Mini prep kit (Qiagen, cat#27106) resulting in the AAV backbone that harbors the multicloning site of MluI-SpeI-EcoR1 (hereby referred to as AAV-MluI-SpeI-EcoR1).

### Inserting variant oligo library into AAV-MPRA backbone

The AAV-MluI-SpeI-EcoR1 plasmid and the variant library were digested with SpeI-HF, MluI-HF, and rSAP for 3 hours at 37°C, and heat inactivated for 20 minutes at 80°C. The digested plasmid (∼4kb) was run through a 1% agarose gel and gel extracted using Zymo Gel DNA Recovery kit (Zymo, cat#D4007). The digested library was cleaned up using Dynabeads MyOne Streptavidin C1 beads (Thermo Fisher, cat#65601). The digested library and plasmid were ligated together using T7 DNA ligase at room temperature for 30 minutes using a 1:3 ratio (plasmid:library). The ligated product was cleaned up using Zymo PCR clean and concentrator-5 (Zymo, cat#11-303C), and transfected into Endura electrocompetent cells via electroporation, and plated on 10 cm circular LB agar plates with ampicillin. The plates were grown overnight at 30°C. The colonies were scraped and grown in 2 L of LB with ampicillin for 7 hours at 37°C. The resulting plasmid was maxi prepped using Qiagen Maxi prep kit (Qiagen, cat#12163) resulting in the AAV library that contains variant-barcode combinations (hereby referred to as an AAV-variant-barcode library).

### Barcode mapping

The variant and barcode region of the AAV-variant-barcode library was PCR amplified using NEBNext 2X Q5 Hifi HS Mastermix with primers that contain Illumina P5 and P7 adapters (*Bcmap_P5_AAV_R* and *Bcmap_P7_AAV_F*). The PCR product was cleaned up using Zymo PCR clean and concentrator-5. The resulting library was sequenced using custom sequencing primers (*BCmap_R1Seq_AAV_R* and *BCmap_R2Seq_AAV_F*) via Novaseq 6000 SP (2×250bp) by the UNC High-Throughput Sequencing Facility (HTSF). Barcodes were assigned to each variant using the custom code available in the github repository (https://github.com/kiminsigne-ucla/bc_map).

### Adding in minimal promoter and GFP

We obtained pLS-minP, a plasmid that contains a minimal promoter (minP) and GFP (minP-GFP), from Dr. Nadav Ahituv’s group (https://www.addgene.org/81225/). The minP-GFP fragment was amplified from the plasmid via PCR using NEBNext 2X Q5 Hifi HS Mastermix and cleaned up using Zymo PCR clean and concentrator-5 (primers: *minP-GFP-F* and *minP-GFP-R*). The minP-GFP fragment and AAV-variant-barcode library were both digested with KpnI-HF (NEB, cat#R3142S) and rSAP for 3 hours at 37°C, which was followed by heat inactivation for 10 minutes at 65°C. Both of these products were then gel extracted from a 0.8% agarose gel using Zymo Gel DNA Recovery kit. The gel extracted products were then digested with XbaI (NEB, cat#R0145S) and rSAP for 3 hours at 37°C, and then for 10 minutes at 65°C for heat inactivation. The digested products were cleaned up using Zymo PCR clean and concentrator-5.

The digested minP-GFP and AAV-variant-barcode library plasmid were ligated together using T7 DNA ligase. The ligation mix was incubated at room temperature for 30 minutes, then cleaned up using Zymo PCR clean and concentrator-5. The ligation mix was transformed into Endura electrocompetent cells, which were then plated on 10 cm circular LB agar plates with ampicillin, resulting in the AAV-variant-minP-GFP-barcode library. The AAV-variant-minP-GFP-barcode library was grown in 2 L of LB with ampicillin, and maxi prepped using Qiagen Maxi prep kit.

The UNC vector core packaged the AAV-variant-minP-GFP-barcode library into AAV serotype 2 (AAV2). The resulting virus had the titer of 7×10^12^ viral particles/uL.

### Administration of AAV-MPRA to HNPs

Acquisition, generation, and culture of human neural progenitors (HNP) have been previously described (Aygün et al., 2021). Donor number 54 was used for all experiments. Briefly, 6-well plates were coated with PBS with Poly-L-Ornithine (10 μg/ml; Sigma-Aldrich, cat#P3655-100MG) and PBS with fibronectin (5 μg/ml; Sigma-Aldrich, cat#F1141-5MG). HNPs were plated at 400K cells/well. The cells were plated in Neurobasal A media (Thermo Fisher, cat#10888022) supplemented with primocin (100 μg/ml; Invitrogen, cat#ant-pm-2), BIT 9500 (10%; STEMCELL, cat#09500), glutamax 100X (1%; Fisher Scientific, cat#5112367), Heparin (1 μg/ml; Sigma-Aldrich, cat#H3393-100KU), and growth factors: EGF (20 μg/ml; PeproTech, cat#AF-100-15), FGF (20 μg/ml; PeproTech, cat#AF-100-15), PDGF (20 ng/ml; PeproTech, cat#100-00AB), and LIF (2 ng/ml; PeproTech, cat# 300-05). The next day, each well was transduced with the AAV-MPRA library at 7,000 multiplicity of infection (MOI). One well per plate was a no-virus control to monitor general cell health. After the AAV-MPRA library was added to the cells, the plates were spun in a centrifuge for 5 minutes at 37°C at 1000 rcf. Cells were half-fed with 2X growth factors every other day for two weeks after transduction. To enhance detectability of transduced cells, we pooled 3 wells for one replicate, resulting in 1.2 million cells per replicate.

### Processing RNA and DNA for sequencing

RNA was extracted from each well using Qiagen RNeasy kit (Qiagen, cat#74004), using 10 uL of *β*-Mercaptoethanol (Sigma-Aldrich, cat#60-24-2) per 1 mL of Qiagen RLT buffer. The columns were treated with DNase (Qiagen, cat#79256). cDNA was generated from the extracted RNA by SuperScript IV Reverse Transcriptase (Invitrogen, cat#18090050) using a primer that targets downstream of the barcodes (*Lib_Hand_RT_AAV*).

To acquire an initial input of DNA put into the cells, DNA was extracted from the AAV2 virus which contained the AAV-MPRA library using a NucleoSpin virus kit (Macherey-Nagel, cat#740983.50).

### Amplification of RNA-seq libraries

DNA extracted from the AAV-MPRA library and cDNA from each transduced well were amplified via PCR using NEBNext 2X Q5 Hifi HS Mastermix (primers for DNA: *Lib_Hand_RT_AAV* and *Lib_Seq_GFP_AAV_R*; primers for cDNA: *Lib_Hand_AAV* and *Lib_Seq_GFP_AAV_R*). The samples were cleaned up using Zymo clean and concentrator-5. This was followed by the second amplification step to add on sequencing adaptors and unique Illumina indices (primers: *P5_Seq_GFP_AAV_F* and *P7_Ind_#_Han*). Again, NEBNext 2X Q5 Hifi HS Mastermix was used for amplification. The resulting libraries were cleaned up using 0.75X ampure beads (Beckman Coulter, cat#A63881) and sequenced by UNC HTSF via Novaseq 6000 SP (1×35bp), with custom primers that capture the barcode sequence and sequencing index (read 1 primer: *Exp_R1_seq_P_AAV*, index primer: *Exp_Ind_seq_P_AAV*).

### Quality check and barcode aggregation

Using the barcode-variant relationship decoded from the barcode mapping step, RNA and DNA barcodes from RNA- and DNA-sequencing were mapped back to their corresponding variants. We counted the number of barcodes mapped to each variant and found that each variant was mapped to ∼200 barcodes on average. Next, we aggregated the RNA and DNA barcode counts for each variant. Because different combinations of barcodes could be introduced to different biological replicates, using the same DNA counts measured from the AAV-MPRA library for all biological replicates could lead to incorrect normalization. To mitigate this, if a given RNA barcode was missing in one biological replicate, that barcode was not counted in aggregating DNA counts for that replicate. This way, even when the DNA from the AAV-MPRA library was used, each biological replicate could have different DNA barcode counts guided by RNA barcodes. For example, if barcodes 1, 2, and 3 were mapped to the variant1, and barcode 2 was missing from the RNA barcode count, we would simply sum up the counts for barcodes 1 and 3 for both DNA and RNA. In contrast, if none of the barcodes were measured, that corresponding variant will have NA count. After merging both DNA and RNA counts by those criteria, we discarded any variants that had more than eight NAs across ten replicates.

### Identification of MPRA-positive variants

Using the aggregated DNA and RNA counts, we used *mpra* Bioconductor package to calculate differential allelic regulatory activity (Myint et al., 2019). We used *mpralm()* function which uses the linear model to measure differential regulatory activity between two alleles with following parameters:

*mpra_lm_object <-mpralm(object = mpra_set, design = design_matrix, aggregate = “none”, normalize = T, block = samples, model_type = “corr_groups”)*

Here, *mpra_set* refers to the *mpra* object created by *MPRAset()* function consisting DNA and RNA counts and *design_matrix* refers to the matrix that specifies the reference and alternative allele status of the corresponding DNA/RNA counts. For our code we used 1) *aggregate = “none”* since we aggregated our barcodes before running *mpralm* and 2) *normalize = T* as DNA and RNA counts were not pre-normalized. Lastly, we named our replicates with the *samples* variable and used *model_type = “corr_groups’’* for paired mixed-model fit.

The resulting *mpra_lm_object* provides summary statistics (e.g. logFC, average expression, t-statistics, P-value, adjusted P-values, and B-statistics) of each variant (**Supplementary Table 1**). We defined MPRA-positive variants as variants that show statistical RNA count difference between reference and alternative allele at FDR < 0.1, while defining MPRA-negative variants as variants with no significant allelic regulatory activity at nominal P > 0.1.

### Measuring reproducibility

We applied *mpralm* normalization method to 10 biological replicates to scale all replicates to have a common size of 10 million reads. We then used *corrplot* R package’s *corrplot.mixed(upper = “number”, lower = “square”, col.lim = c(.5,0.8))* function to compare the RNA/DNA count ratio between biological replicates (**Supplementary Figure 2)**.

### Circular Manhattan Plots

Circular Manhattan plots (**Figure 1B, Supplementary Figure 1)** for finemapped and MPRA variants were created by using *CMplot* R package (Yin et al., 2021). For finemapped GWAS variants, *CMplot(*…, *type = “p”, plot.type = “c”, threshold = 5e-8)* was used. For our MPRA variants, we used threshold = 0.1 and all other parameters were identical.

### LD SNPs and random SNPs

To define the local background, LD SNPs were selected. LD (or a schizophrenia GWS locus) was defined as a region that encompasses SNPs with r^2^>0.6 to the index SNP. All SNPs within 144 schizophrenia GWS loci with nominal association (P<0.001) were selected. Finemapped variants were then extracted from these variants, leaving non-finemapped SNPs with nominal association within LD.

To define the global background, random SNPs with matched minor allele frequency (MAF) and LD were selected. For each finemapped SNP, we randomly selected 10 SNPs within the same chromosome that have matching (±10%) MAF and the number of SNPs in LD (defined as r^2^ > 0.1). If less than 10 SNPs were identified for a given SNP, we selected all SNPs matched with MAF and LD. MAF and the number of LD buddies for genome-wide SNPs were obtained from *garfield* Bioconductor package (Iotchkova et al., 2016).

### Genomic annotation

Using the *annotatr* Bioconductor package, MPRA-positive and MPRA-negative variants were mapped to its corresponding genomic annotations. After labeling MPRA-positive and MPRA-negative variants to its corresponding genomic annotations, we noticed that some of the variants are overlapping with multiple annotations which leads to overrepresentation of certain variants. To mediate this issue, we prioritized certain annotations (i.e., exons/UTRs (can be duplicated) > promoters > 1kb to 5kb from promoter > introns > intergenic) so that each SNP is mapped to a single genomic annotation.

### Epigenetic annotation

We used 1) H3K27ac peaks from the fetal and adult dorsolateral prefrontal cortex (DLPFC) as fetal and adult brain enhancers, respectively (Li et al., 2018), 2) H3K27ac peaks from sorted brain cells as cell-type specific adult brain enhancers (Nott et al., 2019), and 3) single-cell ATAC-seq peaks from the fetal brain as cell-type specific fetal brain enhancers (Ziffra et al., 2021) to compare the epigenetic differences of MPRA-positive, MPRA-negative, LD, and random SNPs. We overlapped MPRA-positive, MPRA-negative, LD, and random SNPs with each enhancer set using *findOverlaps()* function in *GenomicRanges* Bioconductor package. Then their overlap proportion was calculated by dividing the number of overlapped variants by the original number of variants (e.g., 4 out of 10 variants within the variant set A overlapped with enhancer set B gives 40% overlap). To compare the overlap between two SNP categories, Fisher’s exact test with the contingency table below was used.

|  |  |
| --- | --- |
| The number of MPRA-positive variants overlapping with epigenetic region A | The number of MPRA-positive variants not overlapping with epigenetic region A |
| The number of MPRA-negative/LD/random overlapping with epigenetic region A | The number of MPRA-negative/LD/random overlapping with epigenetic region A |

### Evolutionary conservation

From the Zoonomia consortium, we obtained human phyloP scores predicted from the comparative genomic analysis of 240 mammalian species (Zoonomia Consortium, 2020). Since phyloP scores were available in hg38, we converted them to hg19 using liftOver (Navarro Gonzalez et al., 2021). As we used 150 bp sequences centered (76th position) on each variant for our MPRA experiment, we calculated average phyloP scores for 150 bp sequences flanking the variants of interest. Average phyloP scores were calculated for MPRA-positive, MPRA-negative, LD, and random SNPs and compared against each other using Wilcoxon rank sum test. To ensure that this finding is not dependent on the size of the window used, we also used different window sizes (e.g. 100bp, 200bp, and 300bp centered on each variant), but the choice of window sizes did not change the results.

Similar to phyloP scores, average phastCons scores of the same sequences were obtained using the Bioconductor package *phastCons100way.UCSC.hg19* (Siepel et al., 2005).

### TF motif analysis

To observe TF motif altering properties of MPRA-positive variants, we used *motifbreakR* Bioconductor package (Coetzee et al., 2015). Following the *motifbreakR* vignette, we subsetted the TF motif database by *Hsapiens* and excluded *stamlabs* since they are not annotated. This database included TF motif data from *cisbp_1.02, HOCOMOCOv10, HOCOMOCOv11, hDPI, JASPAR_2014, JASPAR_CORE, jaspar2016, jaspar2018, jolma2013, SwissRegulon, and UniPROBE*. Then we ran *motifbreakR(*…, filterp = TRUE, method = “ic”, threshold = 1e-4) and filtered the result by *effect = “strong”* to observe strong TF motif alterations only.

### TF enrichment analysis

To calculate the TF enrichment for MPRA-positive variants, we also ran *motifbreakR* on LD and random SNPs. Then we compared the number of TF motif alterations between MPRA-positive and LD/random SNPs and calculated statistical significance by Fisher’s exact test with the contingency table of

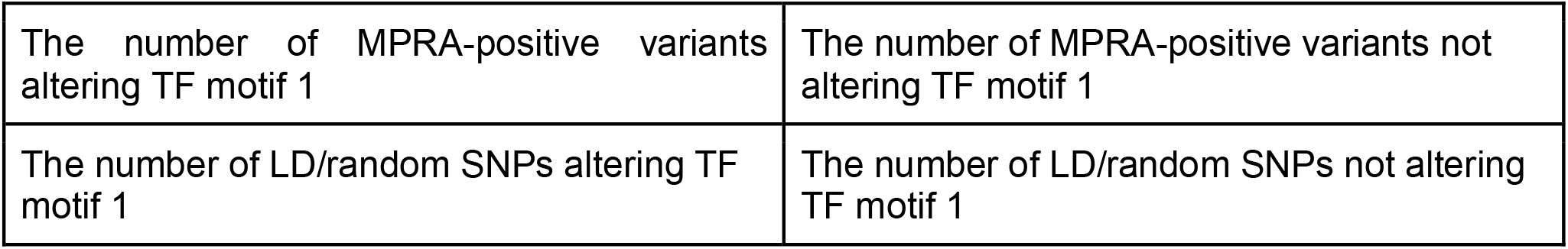

### Calculation of corrected *Δ*SVM scores

To predict the impact of TFs on SNP-mediated regulatory activity, we calculated corrected delta support vector machine (*Δ*SVM) scores with the following formula for each variant.

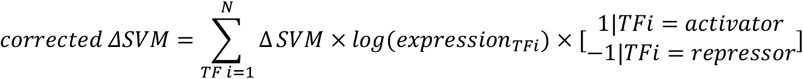

*Δ*SVM scores for each TF-SNP pair were obtained from Yan et al. (Yan et al., 2021); expression levels of TFs in HNPs have been obtained from Aygün et al. (Aygün et al., 2021); information about whether TFs are activators or repressors has been obtained from Savitskaya (Savitskaya, 2010). For TFs that are predicted to act as both activators and repressors, we assumed that they mainly act as an activator.

The resulting corrected *Δ*SVM scores were compared against MPRA logFC values at a variant level. Pearson’s correlation coefficients between corrected *Δ*SVM scores and MPRA logFC were calculated for MPRA-positive and MPRA-negative variants. We then randomly sampled corrected *Δ*SVM scores and MPRA logFC values for MPRA-negative variants for 1,000 times to calculate the permuted distribution of Pearson’s correlation coefficient. The observed Pearson’s correlation coefficient for MPRA-positive variants was compared against the permuted distribution to calculate the permuted P-value.

### eQTL overlap

eQTL datasets from the adult DLPFC (n=1,387) and fetal cortices (n=201) were obtained from Wang et al. (Wang et al., 2018) and Walker et al. (Walker et al., 2019), respectively. We overlapped our MPRA-positive variants with brain eQTL resources by matching variant information (i.e., chromosome, position, rsid). One discrepancy that we found was that our data contained SNPs in chromosome X, whereas both eQTLs lacked SNPs in sex chromosomes.

Colocalization analysis between adult DLPFC eQTLs and schizophrenia GWAS was obtained from Liu et al. (Liu et al., 2021). Same analytic pipeline was used to perform colocalization analysis between developing brain eQTLs and schizophrenia GWAS. Briefly, we intersected developing brain eQTLs with schizophrenia GWS loci using *findOverlap()* function in *GenomicRanges* Bioconductor package. We then performed colocalization analysis between schizophrenia GWAS and eQTLs using the default setting of *coloc* R package (Giambartolomei et al., 2014). We selected loci and eGenes with colocalization posterior probability greater than 0.6 (H4 PP>0.6) to compare against MPRA-positive variants.

For the variant level overlap analysis, the proportion of eQTL overlap was calculated by dividing the number of MPRA-positive variants that overlapped with eQTLs (i.e., matching rsid, chr, and pos) by the total number of MPRA-positive variants. Then, the proportion of IDE overlap was calculated by dividing the number of MPRA-positive-eQTL overlapped variants that has any IDE variant-gene pairs (i.e., MPRA log_2_FC > 0 & eQTL beta > 0 and vice versa) by the number of MPRA-positive-eQTL overlapped variants. Lastly, we overlapped our IDE variants’ genomic coordinates to the colocalized GWS loci using *findOverlap()* function and calculated the overlap by dividing the number of IDE variants that overlapped to the colocalized locus by the number of IDE variants. For each overlap, the number of genes and loci was counted as well.

### TSS distance analysis

Using the Gencode v19 promoter definition (Frankish et al., 2021), we employed *bedtools v 2.29 (Quinlan and Hall, 2010) closest* function to calculate the distance to the nearest promoters for MPRA_non-eQTL_ and MPRA_eQTL_ variants. Then Wilcoxon rank sum test was used to calculate the statistical significance between two distributions.

### Assigning genes to MPRA-positive variants using Hi-C data

To assign genes to MPRA-positive variants using long-range interactome, first we filtered the Hi-C loops from the four datasets (GZ, CP, PN, AN) that interact with Gencode v19 promoters (hereafter referred to as promoter-anchored loops). Then we overlapped 439 MPRA-positive SNP coordinates with the other end of the promoter-anchored loops (the non-promoter anchor) to identify variants that interact with promoters through loops. SNP-gene pairs obtained this way were filtered for protein-coding genes with HUGO Gene Nomenclature Committee (HGNC) symbols, resulting in a total 272 genes (MPRA_Hi-C_ genes). To visualize the loci of MPRA_Hi-C_ genes (variants, genes, Hi-C loops), *plotgardener* Bioconductor package was used (Kramer et al., 2022). When loops were plotted, we only visualized the midpoint of each loop’s end for simplicity.

### Gene ontology

For gene ontology (GO) analysis, we used *gprofiler2* R package (Kolberg et al., 2020). GO terms with term size between 5 and 1000 were filtered, resulting in 26 terms (FDR<0.1). To reduce redundant GO terms, REVIGO web interface was used (http://revigo.irb.hr/).

### LOEUF score

A LOEUF score for each gene was obtained from Karczewski et al. (Karczewski et al., 2020). LOEUF scores for MPRA_eQTL-IDE_ genes were compared against MPRA_Hi-C_ genes. Statistical significance of the difference in LOEUF scores between two gene sets was calculated by the Wilcoxon rank sum test.

### Regulatory complexity

To analyze regulatory complexity, we counted the number of loops anchored at promoters of MPRA_eQTL-IDE_, MPRA_eQTL-IDE_ protein-coding, and MPRA_Hi-C_ genes. Because eQTLs from the adult DLPFC were used to identify MPRA_eQTL-IDE_ and MPRA_eQTL-IDE_ protein-coding genes, we used loops from the adult neuronal Hi-C dataset (Hu et al., 2021). Loops that overlap with the promoter of each gene were selected and counted. Kolmogorov–Smirnov test was used to compare the difference in the number of promoter-anchored loops between MPRA_eQTL-IDE_ and MPRA_Hi-C_ genes.

### Cell type-specific gene expression in fetal and adult prefrontal cortex

In order to visualize cell type-specific gene expression, we used the single-cell gene expression matrix from the fetal (Nowakowski et al., 2017) and adult PFC (Wang et al., 2018). Gene expression matrix was filtered for MPRA_Hi-C_ genes. Then scaled, average expression across all genes was calculated for each cell type as previously described (Sey et al., 2020).

### Adding the chromatin context to allelic activity within multi-variant loci

Multi-variant loci were defined as GWS loci that have more than one MPRA-positive SNP detected. We identified 256 MPRA_Hi-C_ genes that were mapped to the multi-variant loci. To understand how these genes were expressed in schizophrenia, we used transcriptomic signature from postmortem adult brains with schizophrenia (hereby referred to as RNA-seq data) (Gandal et al., 2018). MPRA_Hi-C_ genes whose expression was not detected in RNA-seq data (due to their low expression level) were discarded, leaving 192 genes to compare between MPRA and RNA-seq. Because MPRA logFC values were initially calculated to compare the ratio between alternative and reference alleles, we converted them to compare the ratio between risk and protective alleles. The resulting logFC(risk/protective) values encode disease risk: whether the variant will up- or down-regulate the target gene in schizophrenia. We then aggregated variant-level logFC(risk/protective) values to cognate genes using the following three strategies.

1. Additive model: For each MPRA_Hi-C_ gene, we aggregated logFC(risk/protective) values of all MPRA-positive variants within the GWS locus that were assigned to the gene via Hi-C loops. Using all variants within the GWS locus (regardless of showing chromatin interactions with the gene) gave a similar result.

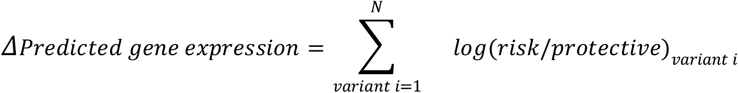
2. Contact model: For each MPRA_Hi-C_ gene, we used all MPRA-positive variants within the locus, because each variant is weighted by contact frequency. We weighted logFC(risk/protective) values with log(normalized contact frequency) between the variant and gene promoter using contact maps of adult neurons (Hu et al., 2021). For a gene with multiple promoters, we used the maximum normalized contact frequency.

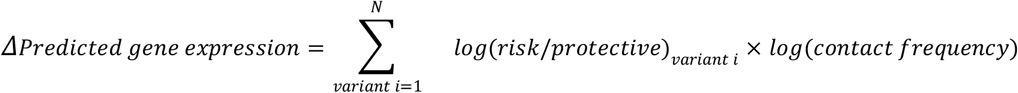
3. Accessibility by contact model: For each MPRA_Hi-C_ gene, we used all MPRA-positive variants within the locus, because each variant is weighted by contact frequency and chromatin accessibility. We weighted logFC(risk/protective) with log(normalized contact frequency) between the variant and gene promoter and average chromatin accessibility of the 150bp element flanking the variant. Contact maps of adult neurons (Hu et al., 2021) and chromatin accessibility from the Brain Open Chromatin Atlas (Fullard et al., 2018) were used to extract contact frequency and chromatin accessibility, respectively. For a gene with multiple promoters, we used the average value of log(normalized contact frequency) x chromatin accessibility.

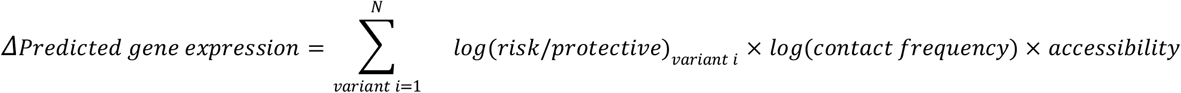

We then compared *Δ*predicted gene expression with RNA-seq logFC values. We did not stratify genes with significant differential expression for this comparison because the effect sizes of common variants are small, which may not necessarily yield significant differential expression in idiopathic schizophrenia. Accordingly, we measured the percentage of genes that show the same direction of effects (e.g. up- or down-regulation) between *Δ*predicted gene expression and RNA-seq logFC.

Because the third model (accessibility by contact model) outperformed other models, we used the same model to calculate *Δ*predicted gene expression from MPRA-negative variants as a control. In addition, we randomly sampled logFC(risk/protective) values for MPRA-positive and - negative variants for 10,000 times to calculate permuted *Δ*predicted gene expression. The percentage of genes that show the same direction of effects between permuted predicted gene expression and RNA-seq logFC was compared against what was predicted from MPRA-positive variants to calculate the permutation P-value.

## Data Availability

Sequencing data are available via the Gene Expression Omnibus under the accession number GSE211045.

## Data Availability

Sequencing data are available via the Gene Expression Omnibus under the accession number GSE211045.

## Code Availability

Custom codes that were used to generate our SCZ MPRA results are available on our GitHub page (https://github.com/thewonlab/schizophrenia-MPRA).

## Contributions

H.W., J.D., and S.K. designed schizophrenia MPRA libraries. J.C.M. created the AAV-MPRA vector and generated MPRA libraries. J.C.M. performed MPRA with help from J.L.B. O.K. helped J.C.M with HNP culture. K.I. created the script to map barcodes to variants. S.L. conducted the bioinformatic analyses of the resulting MPRA datasets. J.L. identified target genes of MPRA-positive variants using Hi-C datasets. M.L.B., D.H.P., and M.I.L. helped establish statistical analytic pipelines. O.K. and J.L.S. optimized transduction protocols for HNPs. H.W., S.L., and J.L. generated figures. H.W., J.C.M., S.L., J.L., and J.L.B. co-wrote the manuscript, which was subsequently reviewed and edited by the rest of the authors.

## Acknowledgement

We thank members of the Won lab for helpful discussions and comments about this paper and Dr. Patrick Sullivan at UNC for his advice on evolutionary conservation analysis. This research was supported by the PsychENCODE consortium (U01MH122509, H.W. and J.L.S.), the IGVF consortium (UM1HG012003, H.W. and M.I.L.), the National Institute of General Medical Sciences (5T32GM067553, S.L; 5T32GM135128, J.C.M. and M.B.), the NIH New Innovator Award from the National Institute of Mental Health (DP2MH122403, H.W.), and the NARSAD Young Investigator Award from the Brain and Behavior Research Foundation (H.W.).

