## Supplementary Figures for "Systematic investigation of allelic regulatory activity of schizophrenia-associated common variants"

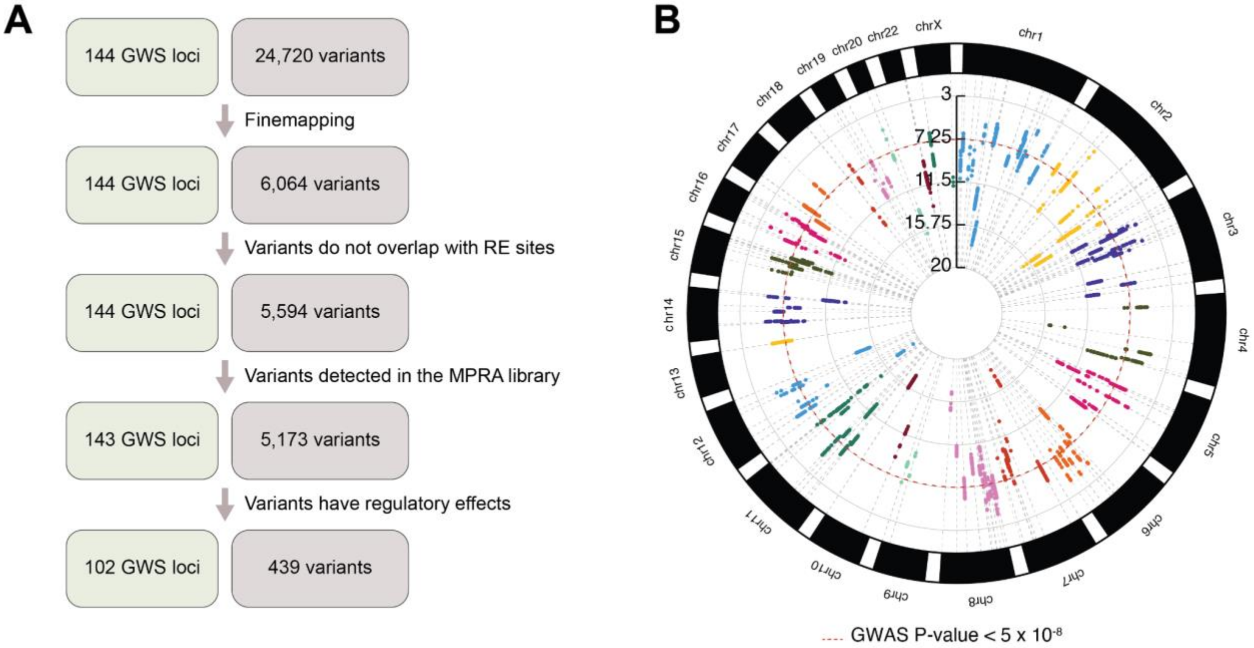


**Supplementary Figure 1. MPRA prioritizes schizophrenia risk variants. A.** A schematic of refining the number of causal variants associated with schizophrenia. In total, 24,720 variants are located in 144 GWS loci associated with schizophrenia at $P<1x10^{-5}$. Computational finemapping identified 6,064 credible variants that can explain the causal configuration of 144 GWS loci. Some of the credible variants were discarded because they overlap with restriction enzyme (RE) sites used for cloning of AAV-MPRA libraries. Some of the variants were missed during the cloning steps, resulting in 5,173 variants to be tested via MPRA. Finally, we identified 439 variants covering 102 GWS loci that show gene regulatory effects. **B.** A circular Manhattan plot displaying the finemapped schizophrenia GWAS variants with their respective P-values. Red dotted line depicts the GWAS P-value threshold of $P<5x10^{-8}$.

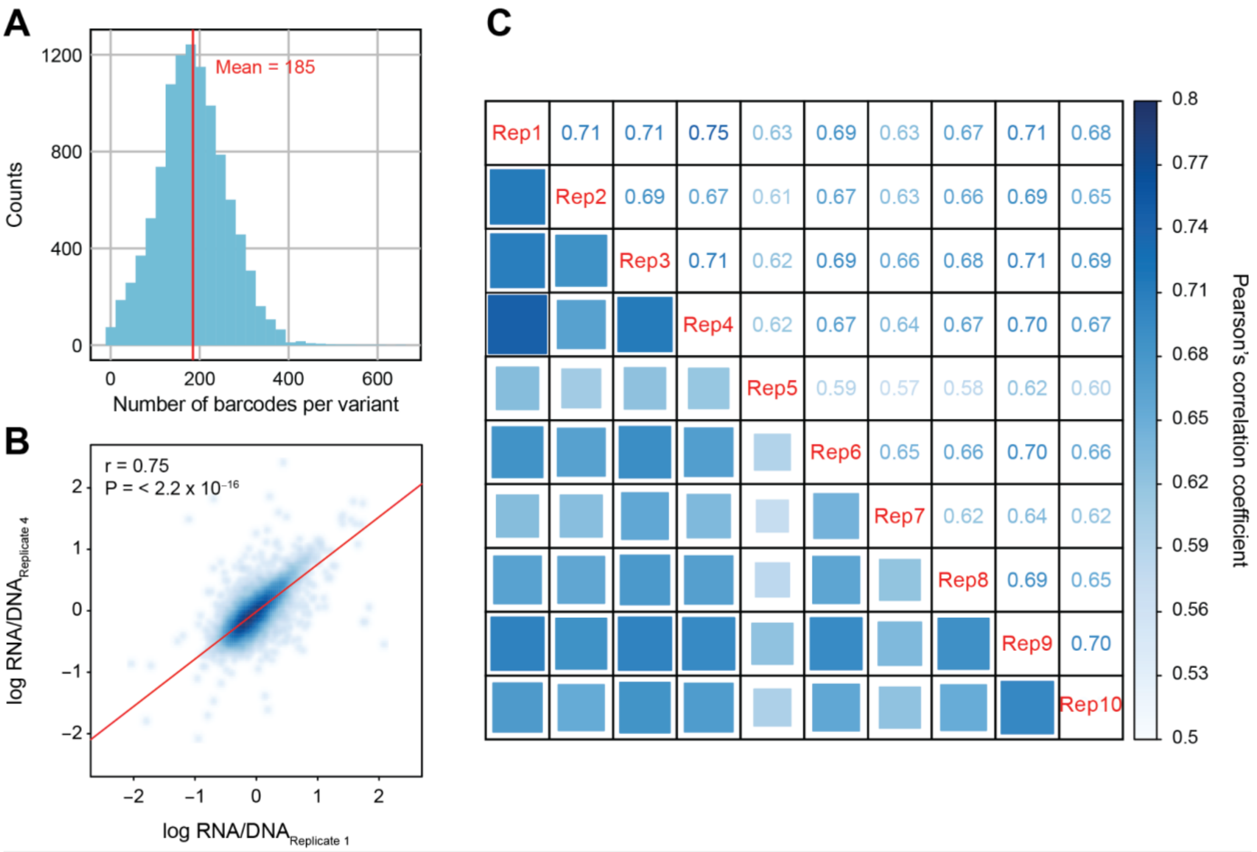


**Supplementary Figure 2. Quality control and reproducibility. A.** The distribution of the number of barcodes assigned to each variant. **B.** Reproducibility between biological replicates 1 and 4. r and P from Pearson's correlation coefficient. **C.** Reproducibility across 10 biological replicates measured by Pearson's correlation coefficients.

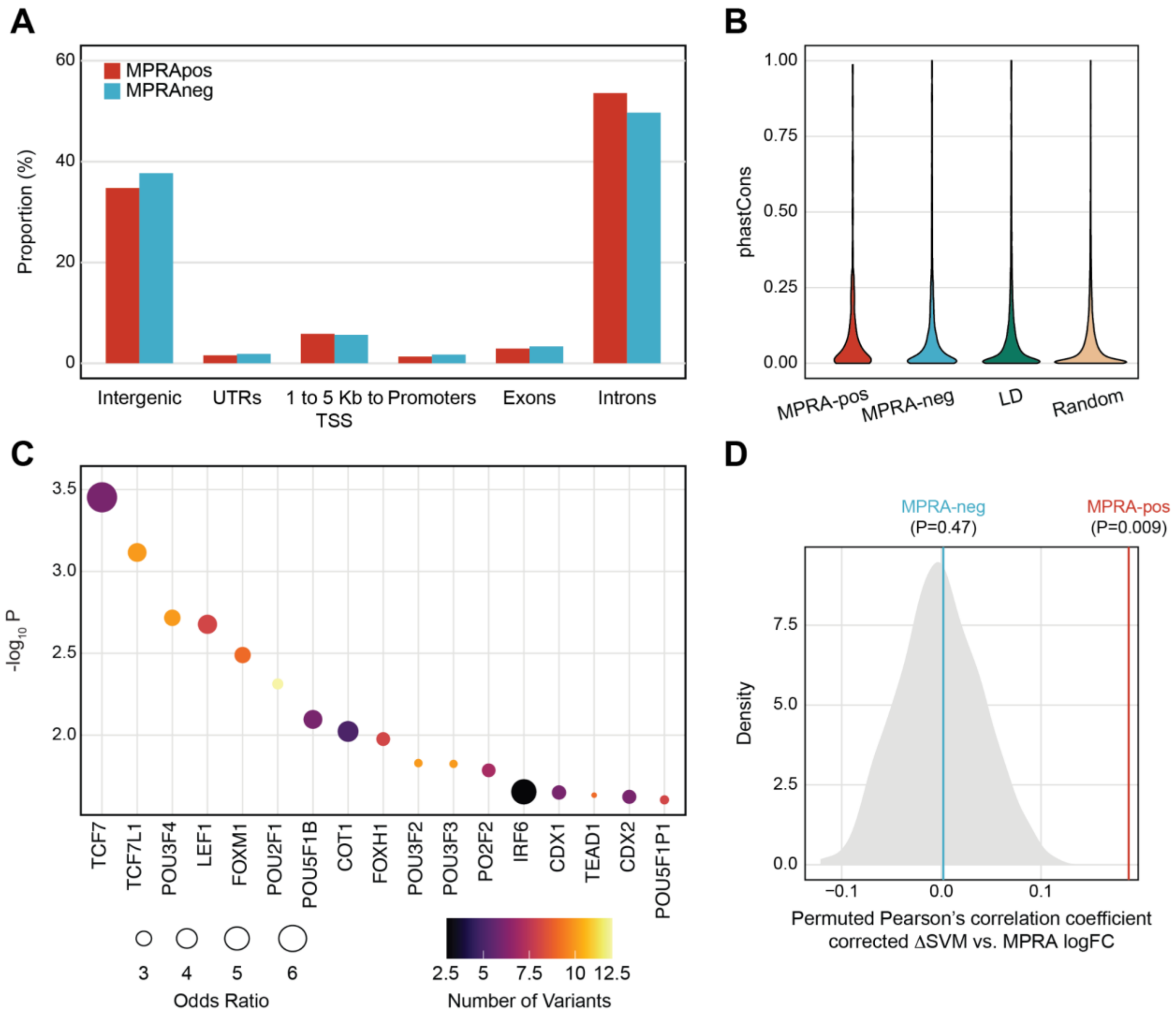


**Supplementary Figure 3. Genomic and epigenomic properties of MPRA-positive variants.** **A.** Genomic annotation of MPRA-positive and -negative variants. UTRs, untranslated regions; TSS, transcription start sites. **B.** PhastCons scores do not differ between MPRA-positive variants and other sets of variants. **C.** TFs whose motifs are predicted to be altered by MPRA-positive variants. TF enrichment was calculated by comparing TF binding motifs between MPRA-positive variants and random SNPs. Each dot is color-coded based on the number of variants that are predicted to alter TF binding motifs. **D.** Pearson's correlation coefficient between TF-mediated regulation (corrected *Δ*SVM) and allelic regulatory activity (MPRA logFC) is significantly higher for MPRA-positive variants than permuted distribution.

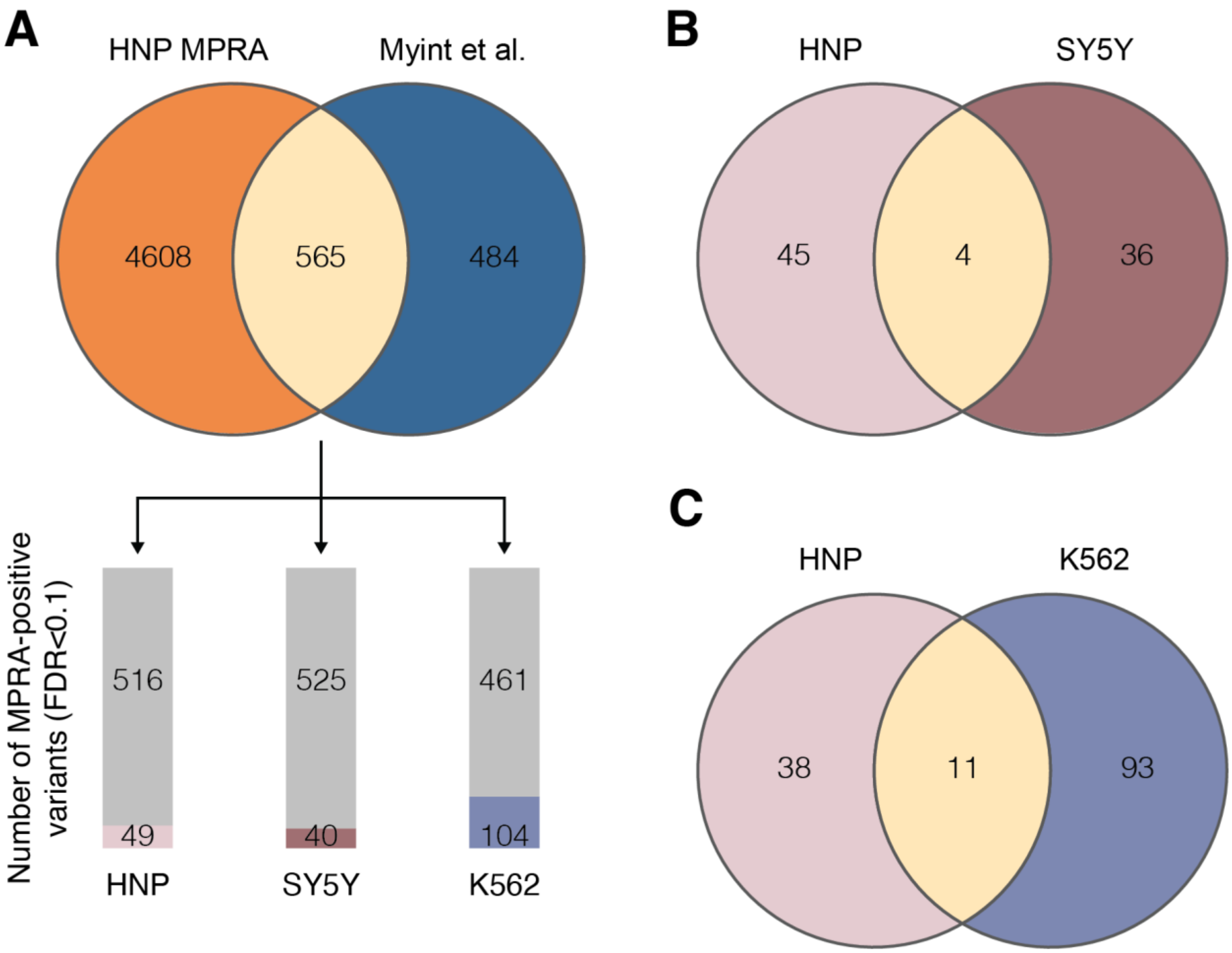


**Supplementary Figure 4. Comparison of MPRA-positive variants defined in different cell types. A.** We compared our MPRA results obtained from HNP (HNP MPRA) with the previously reported MPRA results from SY5Y and K562 (Myint et al.). Top, 565 variants were tested in both studies. Bottom, the number of MPRA-positive variants in HNPs, SY5Y, and K562 at FDR<0.1 among 565 variants tested in both studies. **B.** The overlap of MPRA-positive variants in HNP and SY5Y. **C.** The overlap of MPRA-positive variants in HNP and K562.

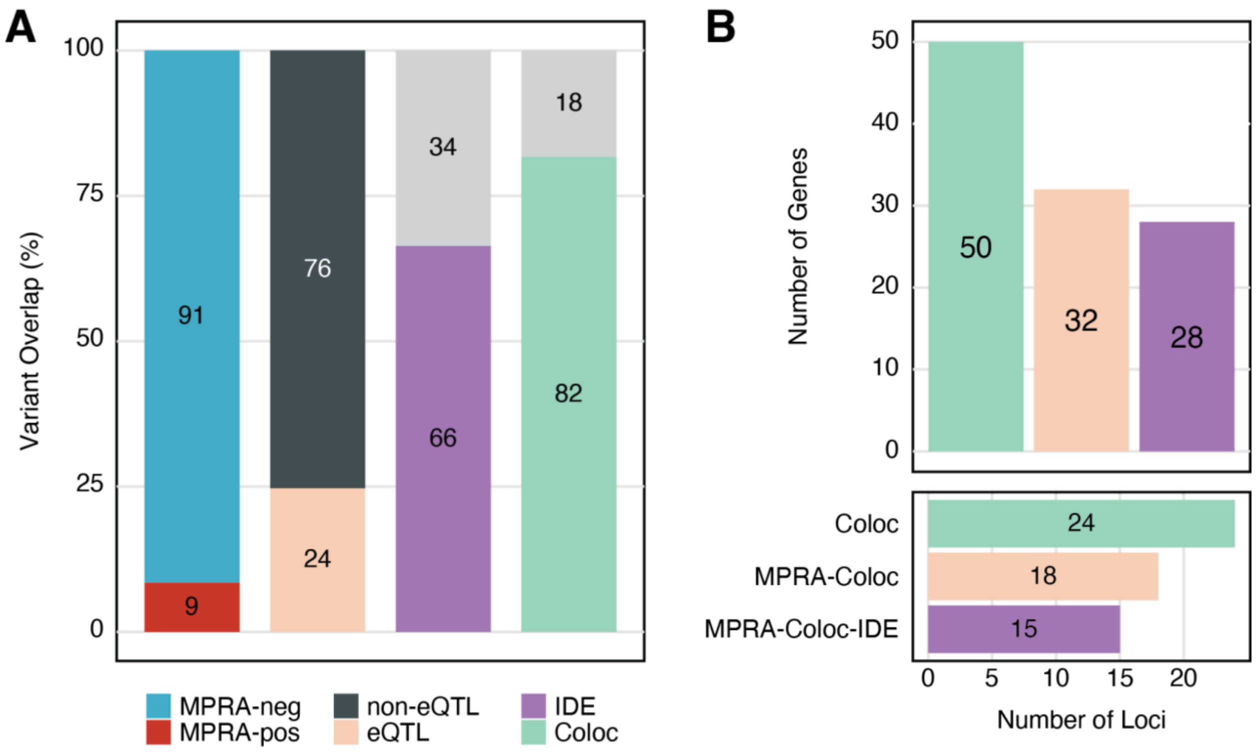


**Supplementary Figure 5. Comparison of MPRA results with developing brain eQTLs. A.** 9% of the variants tested in our MPRA were found to have allelic regulatory effects. 24% of MPRA-positive variants overlapped with developing brain eQTLs. Within the 24% overlap with eQTLs, 66% of MPRA-positive variants are identical in expression direction (IDE) to the overlapping eQTL variants. Within that 66%, 82% of IDE variant-gene pairs were detected from the colocalization analysis between eQTLs and schizophrenia GWAS (Coloc). **B.** 50 schizophrenia GWS loci colocalize with eQTLs, providing 24 schizophrenia-associated eGenes (Coloc). 32 out of these 50 loci contain at least one MPRA-positive variant and are mapped to 18 eGenes (MPRA-Coloc). 28 of MPRA-Coloc loci contain variants that have the identical direction of effects between MPRA and eQTLs and are mapped to 15 eGenes (MPRA-Coloc-IDE).

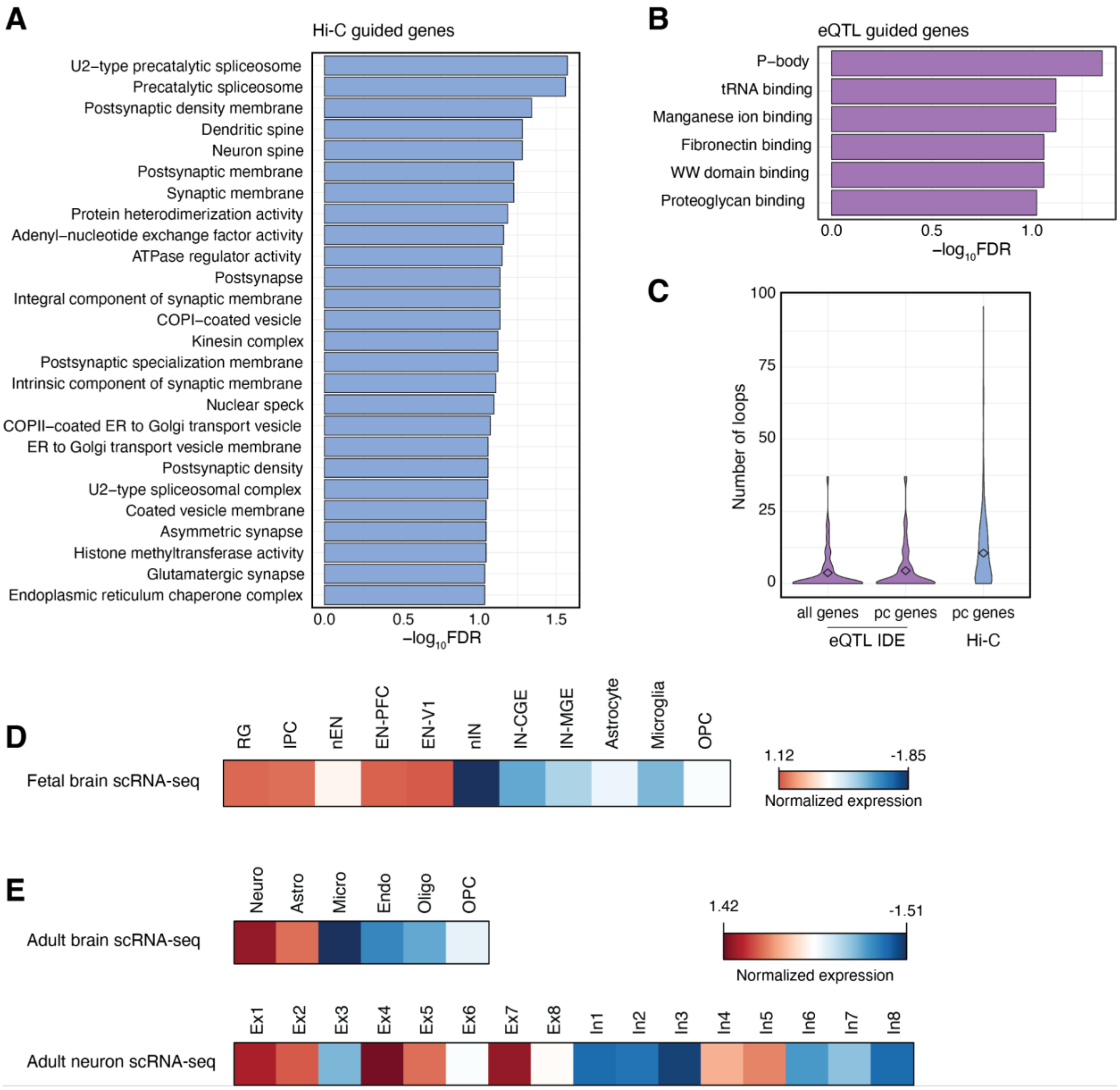


**Supplementary Figure 6. Gene ontology and cell type enrichment of MPRA$_{Hi\text{-}C}$ and MPRA$_{eQTL\text{-}IDE}$ genes A.** Top gene ontology terms for MPRA$_{Hi\text{-}C}$ genes. **B.** Top gene ontology terms for MPRA$_{eQTL\text{-}IDE}$ genes. **C.** Number of promoter-anchored loops for MPRA$_{eQTL\text{-}IDE}$ genes, MPRA$_{eQTL\text{-}IDE}$ protein coding (pc) genes, and MPRA$_{Hi\text{-}C}$ genes. Promoter-anchored loops from adult neurons were used to measure regulatory complexity. **D.** Averaged expression of MPRA$_{Hi\text{-}C}$ genes in different cell types of the fetal brain. RG, radial glia; IPC, intermediate progenitor cells; nEN, neonatal excitatory neuron; EN-PFC, excitatory neuron in the prefrontal cortex; EN-V1, excitatory neuron in the visual cortex; nIN, neonatal inhibitory neuron; IN-CGE, inhibitory neuron in the caudal ganglionic eminence; IN-MGE, inhibitory neuron in the medial ganglionic eminence;

OPC, oligodendrocyte precursor cell. **E.** Averaged expression of $MPRA_{Hi\text{-}C}$ genes in different cell types of the adult brain. Neuro, neuron; Astro, astrocyte; Micro, microglia; Endo, endothelial cell; Oligo, oligodendrocyte; Ex1, L2/3 cortical projecting neuron; Ex2, L3/4 granule neuron; Ex3, L4 granule neuron; Ex4, L4 and L3/5/6 subcortical projecting neuron; Ex5, L4 and L5/6 subcortical projecting neuron; Ex6, L6 neuron; Ex7, L5/6 corticothalamic projecting neuron; Ex8, L6 corticothalamic projecting neuron; In1, VIP+/Reln+/NDNF+ interneuron; In2, VIP+/Reln–/NDNF– interneuron; In3, VIP+/Reln+/NDNF– interneuron; In4, VIP–/Reln+/NDNF+ interneuron; In5, CCK+/nNOS+/Calbindin+ interneuron; In6, Parvalbumin+/CRHBP interneuron; In7, Somatostatin+/Calbindin+/NPY+ interneuron; In8, Somatostatin+/nNOS+ interneuron.

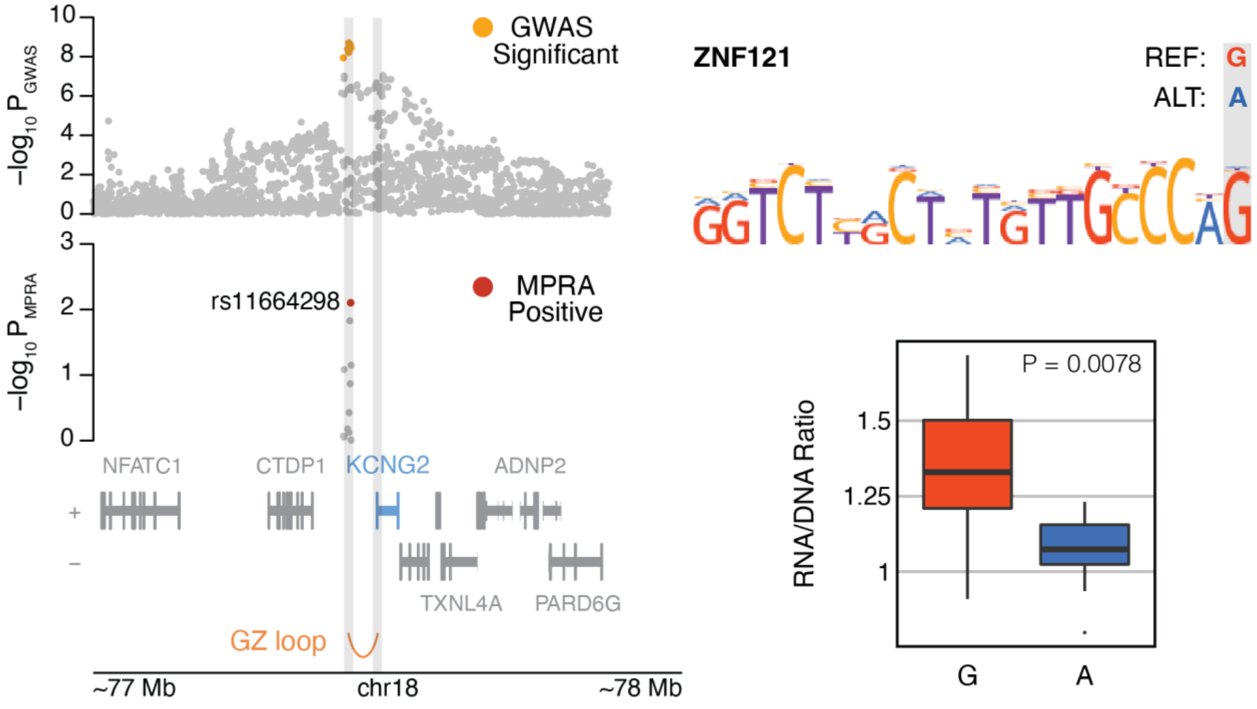


**Supplementary Figure 7. MPRA-positive SNP rs11664298 physically interacts with the promoter of *KCNG2*.** An example locus for *KCNG2* shows that the MPRA-positive SNP rs11664298 physically interacts with *KCNG2* promoter in HNPs. The alternative allele A of rs11664298 breaks the binding motif of ZNF121, which leads to reduced expression of the reporter gene in MPRA.
